# Adrenaline (epinephrine) compared to selective beta-2-agonist in adults or children with acute asthma: a systematic review and meta-analysis

**DOI:** 10.1101/2021.02.17.21251734

**Authors:** Christina Baggott, Jo Hardy, Jenny Sparks, Doñah Sabbagh, Richard Beasley, Mark Weatherall, James Fingleton

**Affiliations:** Medical Research Institute of New Zealand, Wellington, New Zealand; Capital and Coast District Health Board, Wellington, New Zealand; University of Otago Wellington, Wellington, New Zealand

**Keywords:** Asthma, adrenaline, epinephrine, beta_2_-agonists, systematic review, meta-analysis

## Abstract

**Background:** International asthma guidelines recommend against adrenaline administration in acute asthma unless associated with anaphylaxis or angioedema. However, administration of intra-muscular adrenaline in addition to nebulised selective β_2_-agonist is recommended for acute severe or life-threatening asthma in many pre-hospital guidelines. We conducted a systematic review to determine the efficacy of adrenaline in comparison to selective β_2_-agonist in acute asthma.

**Methods:** We included peer-reviewed publications of randomised controlled trials (RCTs) that enrolled children or adults in any healthcare setting and compared adrenaline by any route to selective β_2_-agonist by any route for an acute asthma exacerbation. The primary outcome was treatment failure, as indicated by hospitalisation, stay >24hrs in emergency department, need for intubation, or death.

**Results:** Thirty-eight of 1,140 studies were included, involving 2,275 participants. Overall quality of evidence was low. There was significant statistical heterogeneity, I^2^=56%. The pooled odds ratio for treatment failure with adrenaline versus selective β_2_-agonist was 0.99 (0.74 to 1.34), p=0.96. There was strong evidence that recruitment age-group was associated with different estimates of the risk of treatment failure; with studies recruiting adults-only having a lower risk of treatment failure with adrenaline. It was not possible to determine whether adrenaline in addition to selective β_2_-agonist improved outcomes.

**Conclusion:** The limited evidence available suggests that adrenaline and selective β_2_-agonists have similar efficacy in acute asthma and does not support the use of adrenaline in addition to selective β_2_-agonists in acute asthma. There is a need for high-quality double-blind RCTs to address this issue.

PROSPERO registration number CRD42017079472

## Introduction

Adrenaline, also known as epinephrine, has been used for the management of acute asthma for over 100 years.[1,2] Adrenaline is a non-selective alpha and beta-adrenergic agonist, which causes bronchodilation through stimulation of β_2_ adrenergic receptors leading to relaxation of bronchial smooth muscle. Alpha-agonist effects of adrenaline may also reduce airway oedema.[3,4] Historically, parenteral adrenaline by the subcutaneous or intra-muscular route was the mainstay of acute asthma treatment, [1] but inhaled selective beta2-agonists are now the primary bronchodilators of choice and international asthma guidelines do not recommend the use of adrenaline except in the context of concomitant acute asthma and anaphylaxis or angioedema.[5–8] However, the administration of intra-muscular adrenaline in addition to nebulised selective β_2_-agonist for acute severe or life-threatening asthma remains part of many pre-hospital ambulance guidelines, including in the UK, New Zealand and the USA[9–12] and continues to be used and advocated for in hospital by some clinicians.[4,13–17]

This systematic review and meta-analysis aimed to determine the efficacy of adrenaline in comparison with, and in addition to, selective β_2_-agonist for acute asthma in either children or adults with acute asthma.

## Methods

### Protocol and Registration

This systematic review was prospectively registered on the PROSPERO database on 28th November 2017, registration number CRD42017079472,[18] and is reported in line with the PRISMA guidelines.[19]

### Eligibility Criteria

We included randomised controlled trials that enrolled children, aged 18 months to 17 years, or adults, aged 18 or over, in any healthcare setting, including pre-hospital treatment. All study participants must have had a clinical diagnosis of an acute asthma exacerbation.

Studies in non-English languages were considered for inclusion. The study must have been published in a peer-reviewed journal.

### Types of intervention

The target intervention, as registered on the PROSPERO database, was administration of adrenaline by any route, compared to administration of selective beta-2-agonist by any route.

### Types of outcomes

The primary outcome was treatment failure, as indicated by any one of: hospitalisation, length of stay in emergency department greater than 24 hours, the need for intubation, or death. We recognised *a priori* that these data may not have been reported or able to be extracted from the identified literature. Secondary outcomes of particular interest were peak expiratory flow rate, forced expiratory volume in the first second (FEV_1_), heart rate, length of stay, respiratory rate, oxygen saturation, pH, PCO2, and lactate; as well as the occurrence of adverse events.

### Information Sources and Search

We searched the Cochrane Airways Group Register (Wiley interface, issue 10) and Scopus databases. Our search strategy is detailed in the supplementary appendix and includes search filters for the terms of (‘asthma’ OR ‘asthmatic’) AND (‘epinephrine’ OR ‘adrenaline’). Reference lists from included studies and known reviews were also hand searched. The searches were originally conducted 15^th^ June 2018 and a repeat search on 23^rd^ October 2020 identified no additional relevant studies.

### Study Selection

Selection and data extraction utilised the Covidence online platform.[20] In the screening phase, authors (CB, DS, JH, JF or JS) reviewed the title of the full list of articles identified by the search strategy. After the initial screening, abstracts were examined and articles which appeared to meet the inclusion criteria were reviewed in full by two authors, alongside any articles where there was doubt regarding eligibility. Disagreements were resolved by consensus or arbitration by a third author.

### Data Collection Process and Data items

Data for eligible studies was extracted independently by two authors (CB, JH or JS). Disagreements were resolved by consensus or arbitration by a third author (JF).

### Risk of Bias

We did not plan to examine for publication bias. We assessed reporting bias within individual trials using the risk of bias framework recommended by the Cochrane Collaboration.[21]

### Statistical Analysis

For the primary outcome variable of probability of treatment failure Peto’s method was planned to be used to pool the reported occurrence, to take account of the likely zero cell counts in at least one of the arms. Continuous variables were to be pooled by inverse variance weighting of the mean difference based on reported means, standard deviations, and numbers of participants. Peto’s method was also planned to be used for the occurrence of an adverse event. We planned to evaluate statistical heterogeneity using the I-squared statistic and should it be present to attempt meta-regression based on the following study-level covariates: Route of administration of the intervention, route of administration of the control, predominant age group for the study; and sub-group according to the possible combinations of route of intervention, route of control and whether adrenaline was given in addition to beta-agonist. The method of partitioning the Chi-square statistic was used for the meta-regressions. Both fixed and random effects estimates are shown in the reported pooled analyses.

SAS version 9.4 was used.

## Results

### Study selection

A flow diagram of study selection is shown in Figure 1. A total of 1,140 studies were screened, with 163 undergoing full text review. After full-text review 38 studies were included, involving 2,275 participants. Table 1 provides characteristics of included studies. The route of adrenaline administration was intramuscular for one study, nebulised in six and subcutaneous in the remaining 31. Two studies contained an arm in which adrenaline was administered in addition to selective β_2_-agonist, and in a further 5 studies some participants crossed over to receive adrenaline having previously received selective β_2_-agonist. In the remainder the group receiving adrenaline did not receive selective β_2_-agonist.

**Table 1:**
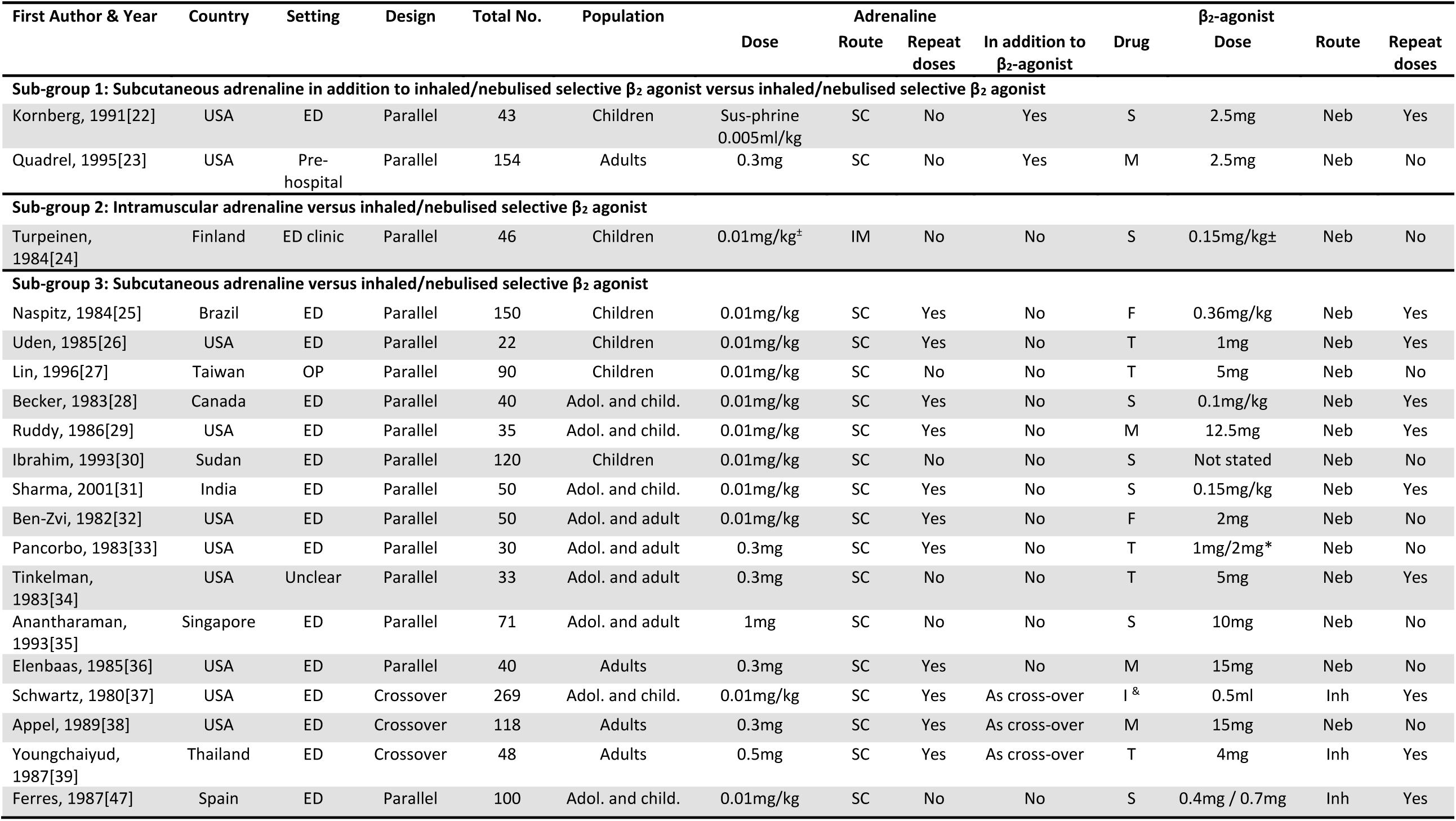
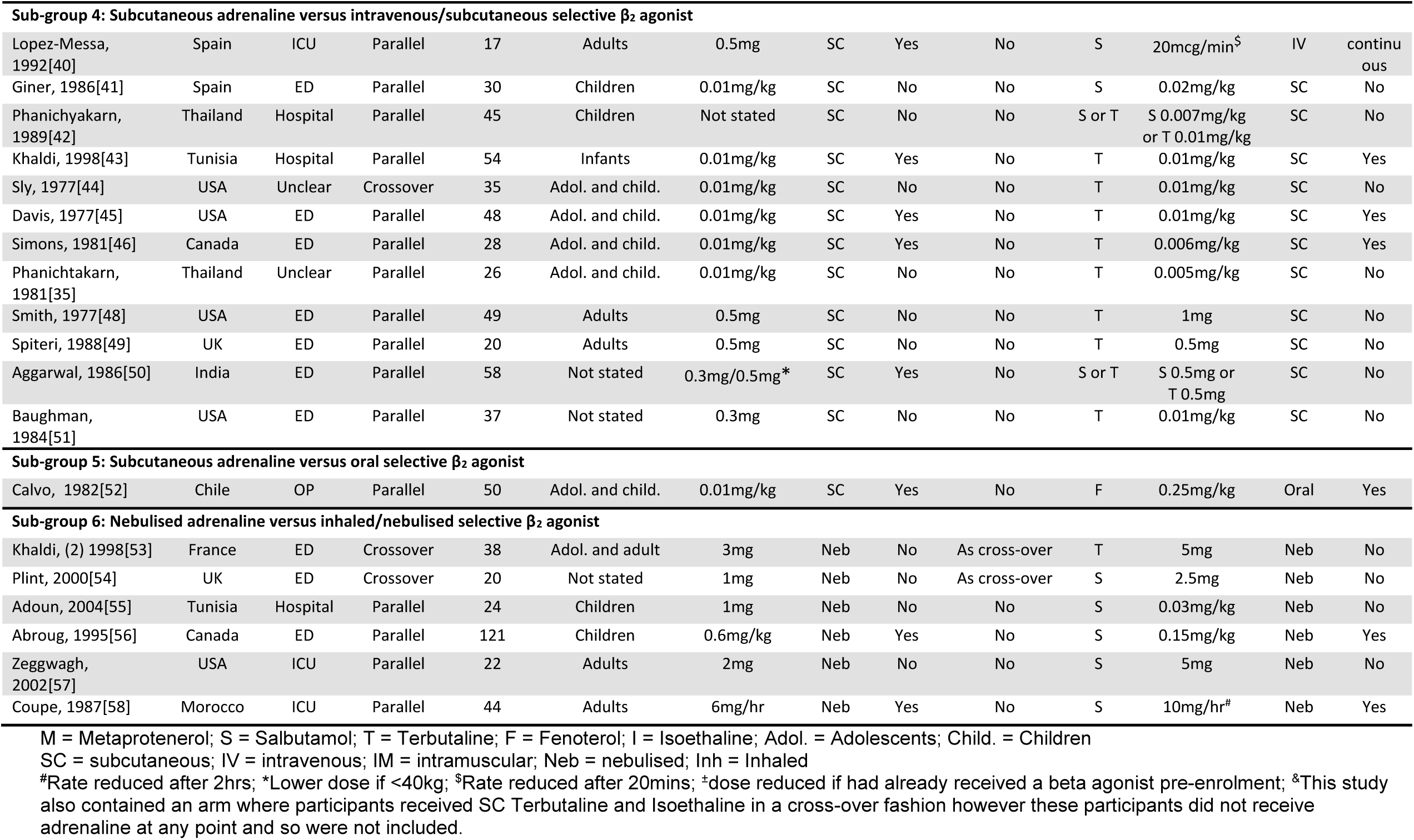
Characteristics of included studies.

| First Author & Year | Country | Setting | Design | Total No. | Population | Dose | Adrenaline<br>Route | Repeat<br>doses | In addition to<br>$\beta_2$-agonist | Drug | $\beta_2$-agonist<br>Dose | Route | Repeat<br>doses |
| --- | --- | --- | --- | --- | --- | --- | --- | --- | --- | --- | --- | --- | --- |
| **Sub-group 1: Subcutaneous adrenaline in addition to inhaled/nebulised selective $\beta_2$ agonist versus inhaled/nebulised selective $\beta_2$ agonist** |  |  |  |  |  |  |  |  |  |  |  |  |  |
| Kornberg, 1991[22] | USA | ED | Parallel | 43 | Children | Sus-phrine<br>0.005ml/kg | SC | No | Yes | S | 2.5mg | Neb | Yes |
| Quadrel, 1995[23] | USA | Pre-hospital | Parallel | 154 | Adults | 0.3mg | SC | No | Yes | M | 2.5mg | Neb | No |
| **Sub-group 2: Intramuscular adrenaline versus inhaled/nebulised selective $\beta_2$ agonist** |  |  |  |  |  |  |  |  |  |  |  |  |  |
| Turpeinen, 1984[24] | Finland | ED clinic | Parallel | 46 | Children | 0.01mg/kg<sup>±</sup> | IM | No | No | S | 0.15mg/kg<sup>±</sup> | Neb | No |
| **Sub-group 3: Subcutaneous adrenaline versus inhaled/nebulised selective $\beta_2$ agonist** |  |  |  |  |  |  |  |  |  |  |  |  |  |
| Naspitz, 1984[25] | Brazil | ED | Parallel | 150 | Children | 0.01mg/kg | SC | Yes | No | F | 0.36mg/kg | Neb | Yes |
| Uden, 1985[26] | USA | ED | Parallel | 22 | Children | 0.01mg/kg | SC | Yes | No | T | 1mg | Neb | Yes |
| Lin, 1996[27] | Taiwan | OP | Parallel | 90 | Children | 0.01mg/kg | SC | No | No | T | 5mg | Neb | No |
| Becker, 1983[28] | Canada | ED | Parallel | 40 | Adol. and child. | 0.01mg/kg | SC | Yes | No | S | 0.1mg/kg | Neb | Yes |
| Ruddy, 1986[29] | USA | ED | Parallel | 35 | Adol. and child. | 0.01mg/kg | SC | Yes | No | M | 12.5mg | Neb | Yes |
| Ibrahim, 1993[30] | Sudan | ED | Parallel | 120 | Children | 0.01mg/kg | SC | No | No | S | Not stated | Neb | No |
| Sharma, 2001[31] | India | ED | Parallel | 50 | Adol. and child. | 0.01mg/kg | SC | Yes | No | S | 0.15mg/kg | Neb | Yes |
| Ben-Zvi, 1982[32] | USA | ED | Parallel | 50 | Adol. and adult | 0.01mg/kg | SC | Yes | No | F | 2mg | Neb | No |
| Pancorbo, 1983[33] | USA | ED | Parallel | 30 | Adol. and adult | 0.3mg | SC | Yes | No | T | 1mg/2mg* | Neb | No |
| Tinkelman, 1983[34] | USA | Unclear | Parallel | 33 | Adol. and adult | 0.3mg | SC | No | No | T | 5mg | Neb | Yes |
| Anantharaman, 1993[35] | Singapore | ED | Parallel | 71 | Adol. and adult | 1mg | SC | No | No | S | 10mg | Neb | No |
| Elenbaas, 1985[36] | USA | ED | Parallel | 40 | Adults | 0.3mg | SC | Yes | No | M | 15mg | Neb | No |
| Schwartz, 1980[37] | USA | ED | Crossover | 269 | Adol. and child. | 0.01mg/kg | SC | Yes | As cross-over | I & | 0.5ml | Inh | Yes |
| Appel, 1989[38] | USA | ED | Crossover | 118 | Adults | 0.3mg | SC | Yes | As cross-over | M | 15mg | Neb | No |
| Youngchaiyud, 1987[39] | Thailand | ED | Crossover | 48 | Adults | 0.5mg | SC | Yes | As cross-over | T | 4mg | Inh | Yes |
| Ferres, 1987[47] | Spain | ED | Parallel | 100 | Adol. and child. | 0.01mg/kg | SC | No | No | S | 0.4mg / 0.7mg | Inh | Yes |
| **Sub-group 4: Subcutaneous adrenaline versus intravenous/subcutaneous selective $\beta_2$ agonist** |  |  |  |  |  |  |  |  |  |  |  |  |  |
| Lopez-Messa, 1992[40] | Spain | ICU | Parallel | 17 | Adults | 0.5mg | SC | Yes | No | S | 20mcg/min<sup>\$</sup> | IV | continuous |
| Giner, 1986[41] | Spain | ED | Parallel | 30 | Children | 0.01mg/kg | SC | No | No | S | 0.02mg/kg | SC | No |
| Phanichyakarn, 1989[42] | Thailand | Hospital | Parallel | 45 | Children | Not stated | SC | No | No | S or T | S 0.007mg/kg or T 0.01mg/kg | SC | No |
| Khalidi, 1998[43] | Tunisia | Hospital | Parallel | 54 | Infants | 0.01mg/kg | SC | Yes | No | T | 0.01mg/kg | SC | Yes |
| Sly, 1977[44] | USA | Unclear | Crossover | 35 | Adol. and child. | 0.01mg/kg | SC | No | No | T | 0.01mg/kg | SC | No |
| Davis, 1977[45] | USA | ED | Parallel | 48 | Adol. and child. | 0.01mg/kg | SC | Yes | No | T | 0.01mg/kg | SC | Yes |
| Simons, 1981[46] | Canada | ED | Parallel | 28 | Adol. and child. | 0.01mg/kg | SC | Yes | No | T | 0.006mg/kg | SC | Yes |
| Phanichtakarn, 1981[35] | Thailand | Unclear | Parallel | 26 | Adol. and child. | 0.01mg/kg | SC | No | No | T | 0.005mg/kg | SC | No |
| Smith, 1977[48] | USA | ED | Parallel | 49 | Adults | 0.5mg | SC | No | No | T | 1mg | SC | No |
| Spiteri, 1988[49] | UK | ED | Parallel | 20 | Adults | 0.5mg | SC | No | No | T | 0.5mg | SC | No |
| Aggarwal, 1986[50] | India | ED | Parallel | 58 | Not stated | 0.3mg/0.5mg* | SC | Yes | No | S or T | S 0.5mg or T 0.5mg | SC | No |
| Baughman, 1984[51] | USA | ED | Parallel | 37 | Not stated | 0.3mg | SC | No | No | T | 0.01mg/kg | SC | No |
| **Sub-group 5: Subcutaneous adrenaline versus oral selective $\beta_2$ agonist** |  |  |  |  |  |  |  |  |  |  |  |  |  |
| Calvo, 1982[52] | Chile | OP | Parallel | 50 | Adol. and child. | 0.01mg/kg | SC | Yes | No | F | 0.25mg/kg | Oral | Yes |
| **Sub-group 6: Nebulised adrenaline versus inhaled/nebulised selective $\beta_2$ agonist** |  |  |  |  |  |  |  |  |  |  |  |  |  |
| Khalidi, (2) 1998[53] | France | ED | Crossover | 38 | Adol. and adult | 3mg | Neb | No | As cross-over | T | 5mg | Neb | No |
| Plint, 2000[54] | UK | ED | Crossover | 20 | Not stated | 1mg | Neb | No | As cross-over | S | 2.5mg | Neb | No |
| Adoun, 2004[55] | Tunisia | Hospital | Parallel | 24 | Children | 1mg | Neb | No | No | S | 0.03mg/kg | Neb | No |
| Abroug, 1995[56] | Canada | ED | Parallel | 121 | Children | 0.6mg/kg | Neb | Yes | No | S | 0.15mg/kg | Neb | Yes |
| Zeggwagh, 2002[57] | USA | ICU | Parallel | 22 | Adults | 2mg | Neb | No | No | S | 5mg | Neb | No |
| Coupe, 1987[58] | Morocco | ICU | Parallel | 44 | Adults | 6mg/hr | Neb | Yes | No | S | 10mg/hr<sup>#</sup> | Neb | Yes |

M = Metaprotenerol; S = Salbutamol; T = Terbutaline; F = Fenoterol; I = Isoethaline; Adol. = Adolescents; Child. = Children
SC = subcutaneous; IV = intravenous; IM = intramuscular; Neb = nebulised; Inh = Inhaled
<sup>#</sup>Rate reduced after 2hrs; \*Lower dose if <40kg; <sup>\$</sup>Rate reduced after 20mins; <sup>\*</sup>dose reduced if had already received a beta agonist pre-enrolment; <sup>&</sup>This study also contained an arm where participants received SC Terbutaline and Isoethaline in a cross-over fashion however these participants did not receive adrenaline at any point and so were not included.

**Figure 1:**
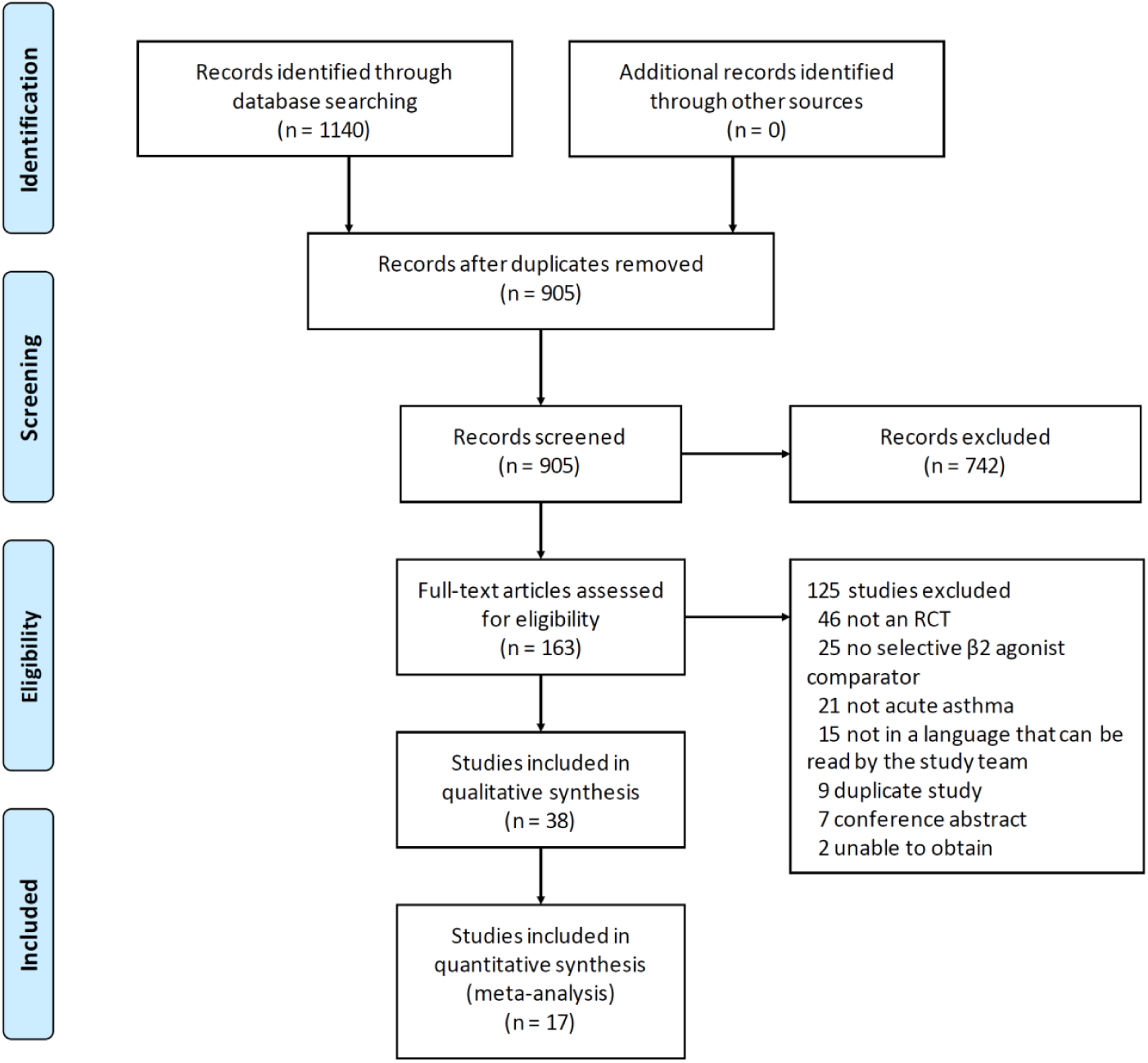
PRISMA flow diagram.

### Risk of bias within studies

Risk of bias assessments are shown in Figure 2. The most recent study was conducted 15 years before this review and 26/38 were conducted more than 30 years prior. Manuscript style, methodology and reporting reflect the norms applicable at the time of publication. As a result, details of study methodology and primary outcome variables were unclear in many cases and 27/38 studies were adjudged to be at high risk of bias in at least one aspect.

**Figure 2:**
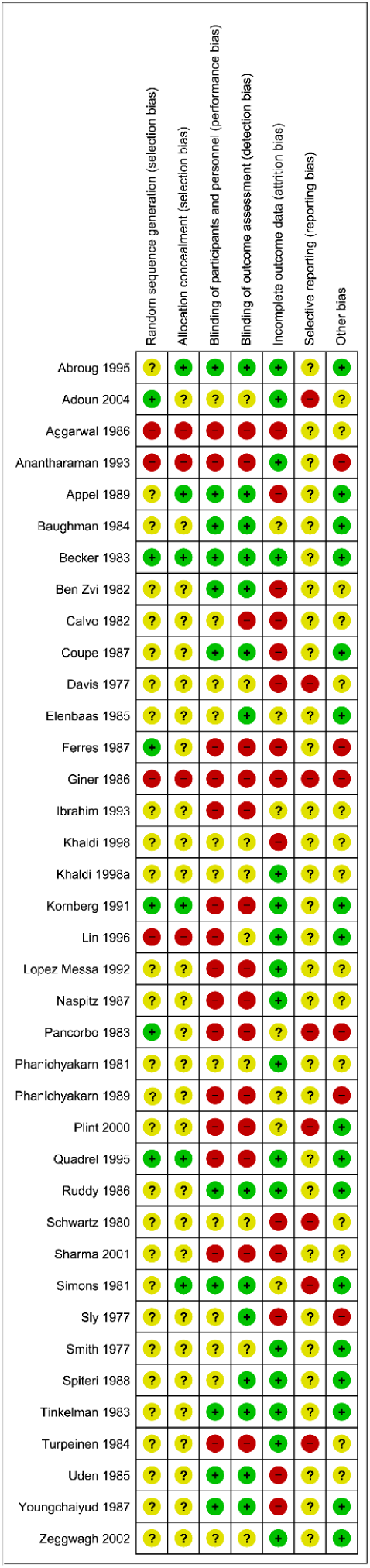

### Synthesis of results

Data on treatment failure was available for 17/38 studies. The individual and pooled study estimates are shown in Table 2 and illustrated in the Forest plot in Figure 3. There was important statistical heterogeneity with a Chi-square statistic of 36.4, (16 DF), P=0.003;, I^2^ (95% CI) 56% (24.4 to 74.5). Neither the fixed or random effects estimates favoured one or other treatment: Peto’s odds ratio pooled fixed effect 0.99 (0.74 to 1.34), p=0.96; and random effects 1.07 (0.65 to 1.76), p=0.79.

**Table 2:** Counts and odds ratio for treatment failure in included studies.

| First Author & Year | Treatment failure<br>n/N (%) |  | Peto's OR (95% CI) | P |
| --- | --- | --- | --- | --- |
|  | Adrenaline | Other |  |  |
| Adoun 2004 | 4/18 (22.2) | 9/20 (45) | 0.37 (0.1 to 1.4) | 0.14 |
| Anantharaman 1993 | 4/27 (14.8) | 6/17 (35.3) | 0.32 (0.08 to 1.34) | 0.12 |
| Appel 1989 | 6/54 (11.1) | 18/46 (39.1) | 0.22 (0.09 to 0.55) | 0.001 |
| Ben-Zvi 1982 | 4/24 (16.7) | 0/26 (0) | 9.20 (1.21 to 69.7) | 0.032 |
| Giner 1986 | 5/15 (33.3) | 3/15 (20) | 1.93 (0.39 to 9.49) | 0.42 |
| Kornberg 1991 | 1/20 (5) | 3/23 (13) | 0.39 (0.05 to 3.03) | 0.37 |
| Naspitz 1984 | 32/75 (42.7) | 22/75 (29.3) | 1.78 (0.91 to 3.45) | 0.09 |
| Pancorbo 1983 | 4/15 (26.7) | 5/15 (33.3) | 0.74 (0.16 to 3.42) | 0.70 |
| Plint 2000 | 9/60 (15) | 8/62 (12.9) | 1.19 (0.43 to 3.30) | 0.74 |
| Quadrel 1995 | 1/53 (1.9) | 1/49 (2) | 0.92 (0.06 to 15.01) | 0.96 |
| Ruddy 1986 | 4/16 (25) | 1/20 (5) | 5.08 (0.78 to 33.12) | 0.09 |
| Schwartz 1980 | 15/66 (22.7) | 58/203 (28.6) | 0.74 (0.4 to 1.39) | 0.35 |
| Sharma 2001 | 4/25 (16) | 4/25 (16) | 1.00 (0.22 to 4.47) | 0.99 |
| Sly 1977 | 5/20 (25) | 1/20 (5) | 4.62 (0.83 to 25.6) | 0.08 |
| Tinkelman 1983 | 0/18 (0) | 1/15 (6.7) | 0.11 (0 to 5.68) | 0.27 |
| Turpeinen 1984 | 9/20 (45) | 5/26 (19.2) | 3.29 (0.94 to 11.52) | 0.063 |
| Uden 1985 | 1/12 (8.3) | 0/7 (0) | 4.87 (0.08 to 283.3) | 0.45 |
| Pooled Fixed effect |  |  | 0.99 (0.74 to 1.34) | 0.96 |
| Pooled Random effect |  |  | 1.07 (0.65 to 1.76) | 0.79 |

**Figure 3:**
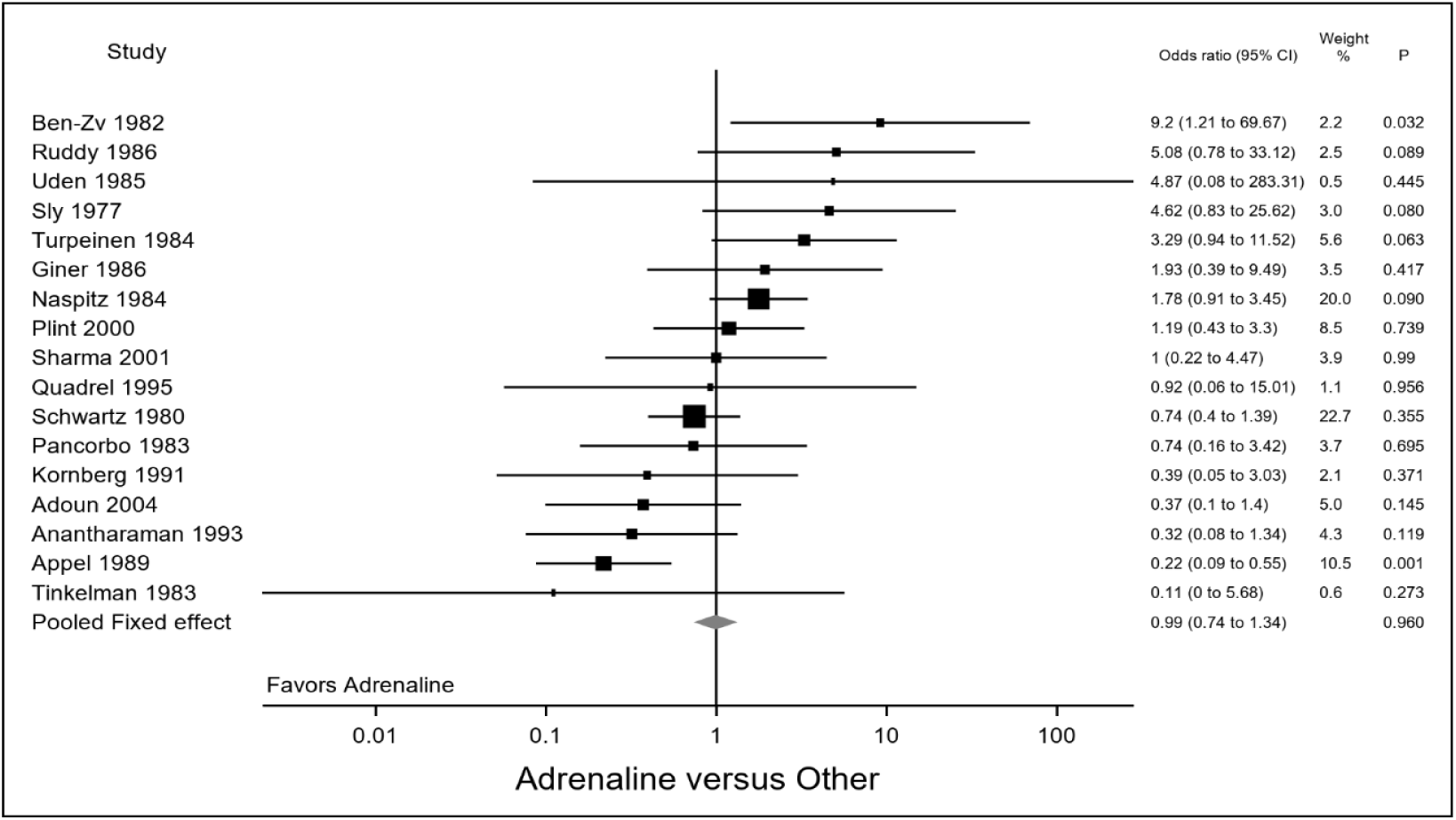
Meta-analysis of odds of treatment failure for adrenaline by any route versus selective β_2_agonist by any route.

For the four sets of study-level covariates there was no evidence that heterogeneity was explained by the intervention route of administration (Table 3). There was weak evidence (P=0.06) that some of the heterogeneity was explained by the control route of administration with the parenteral route at higher risk of treatment failure compared to the inhaled route; although for the latter there was still important heterogeneity within that sub-group. There was weak evidence (P=0.09) that some of the heterogeneity was explained by the sub-groups with sub-groups 2 (IM adrenaline versus nebulised selective β_2_-agonist) and 4 (SC adrenaline versus parenteral selective β_2_-agonist) having a higher risk of treatment failure than the other three sub-groups; although there was still important heterogeneity within the sub-groups. Finally there was strong evidence that the recruitment age-group was associated with different estimates of the risk of treatment failure (P=0.006); with the studies recruiting adults-only having a lower risk of treatment failure with parenteral adrenaline and children-only a higher risk of treatment failure. There was still unexplained heterogeneity within studies that recruited a mixture of adults and adolescents.

**Table 3:** Meta-regression and sub-group effects odds ratios for treatment failure.

| **Study-level covariate** | **Fixed effects Peto odds ratio (95% CI)** | **Between-groups Chi-square statistic (DF)** |
| --- | --- | --- |
| **Intervention route of administration** |  | 3.94 (2)<br>P=0.14 |
| Intramuscular N=1 | 3.29 (0.94 to 11.5) |  |
| Nebulized N=2 | 0.77 (0.34 to 1.73) |  |
| Subcutaneous N=14 | 0.95 (0.68 to 1.33) |  |
| **Control route of administration** |  | 3.55 (1)<br>P=0.06 |
| Inhaled N=15 | 0.92 (0.68 to 1.25) |  |
| Parenteral N=2 | 2.89 (0.90 to 9.28) |  |
| **Sub-group** |  | 8.1 (4)<br>P=0.09 |
| 1 N=2 | 0.53 (0.10 to 2.75) |  |
| 2 N=1 | 3.3 (0.94 to 11.5) |  |
| 3 N=10 | 0.88 (0.62 to 1.27) |  |
| 4 N=2 | 2.89 (0.90 to 9.28) |  |
| 5 N=0 | - |  |
| 6 N=2 | 0.77 (0.34 to 1.73) |  |
| **Overall** |  | **Total heterogeneity Chi-square statistic (DF)** |
| **Total studies** | 0.99 (0.74 to 1.34) | 36.4 (16)<br>P=0.003 |
|  |  | **Between-groups Chi-square statistic (DF)** |
| **Age-group<sup>1</sup>** |  | 12.4 (3)<br>P=0.006 |
| Adults and Adolescents N=4 | 0.79 (0.32 to 2.0) |  |
| Adults and Children N=4 | 1.06 (0.63 to 1.80) |  |
| Adults only N=2 | 0.25 (0.11 to 0.60) |  |
| Children only N=6 | 1.48 (0.91 to 2.41) |  |
| **Overall** |  | **Total heterogeneity Chi-square statistic (DF)** |
| **Total studies**<br>(without Plint, 2000) | 0.98 (0.72 to 1.33) | 36.3 (15)<br>P=0.002 |

1. Plint, 2000 didn't specify age-group and so was not entered into this analysis
The meta-regression by the Chi-square method splits the total heterogeneity Chi-square (with degrees of freedom equal to the total number of studies minus 1) statistic into two components. The between-groups component refers to how much heterogeneity is explained by having a number of study-level covariate groupings, with degrees of freedom of the number of groupings minus 1. Not shown in the table is the within-groups component which is calculated by simple subtraction of total minus within-groups, and reflects how much heterogeneity is still present within the study-level covariate groupings. Where the P value for within-groups heterogeneity is small this reflects evidence that overall heterogeneity is explained, at least in part, by the sub-groups; and is illustrated by the within-group pooled estimates compared to the overall fixed effects estimate without sub-groups.

There was also marked heterogeneity in design, country, population, intervention and control between studies. It was usually not possible to extract secondary outcome data. As a result combining secondary outcome data for meta-analysis was inappropriate and a narrative summary of these data was performed. For this summary, because of the heterogeneity of study design, the studies have been grouped according to the nature of the intervention and comparator. Summary details of key outcomes and adverse events are given in Tables S1 and S2 respectively in the supplementary appendix.

### Sub-group 1: Subcutaneous adrenaline plus nebulised selective β_2_-agonist compared to nebulised selective β2-agonist alone

The question most closely linked to current clinical practice is whether addition of IM adrenaline to inhaled or nebulised selective β_2_-agonist improves outcome. No study addressed this issue. Two of the 38 studies included arms where one group received subcutaneous adrenaline in addition to selective β_2_-agonist and another received selective β_2_-agonist alone.

Kornberg et al. reported an unblinded parallel group RCT of 43 children aged 3 to 12yrs attending a paediatric emergency department for acute asthma. Patients received either 2.5mg salbutamol nebuliser repeated as required every 20 to 30 minutes or a single subcutaneous injection of Sus-Phrine (a combination of immediate- and sustained-release forms of adrenaline) 0.005ml/kg followed by 2.5mg salbutamol nebuliser repeated as required every 20 to 30 minutes. Treatment response was assessed at 20 and 120min. Treatment failure, in the form of hospitalisation, was present in 1/20 patients in the adrenaline group compared with 3/23 patients who received only selective β_2_-agonist. Both groups showed significant improvement in clinical score, peak flow and respiratory rate; with no difference between the groups.

Quadrel et al reported an unblinded three arm parallel group RCT of 154 adult patients receiving pre-hospital treatment for acute asthma. The authors compared nebulised metaprotenerol 2.5mg, subcutaneous adrenaline 0.3mg, or both. Peak flow improved with treatment in all three groups and there was no difference in treatment failure, peak flow, respiratory rate or heart rate between the groups.

### Sub-groups 2&3: Parenteral adrenaline compared to nebulised selective β_2_-agonist

Seventeen studies compared parenteral (IM or SC) adrenaline with nebulised selective β_2_-agonist. Ten studies reported no difference in efficacy as measured by either treatment failure or lung function. Five studies suggested that selective β_2_-agonist was a more effective bronchodilator and two studies suggested adrenaline was more effective. There was weak and inconsistent evidence suggesting more frequent side-effects with adrenaline. Six studies reported more frequent side-effects (including tremor, agitation and headache) with adrenaline, two studies reported more frequent tremor with selective β_2_-agonist (both used terbutaline), five studies reported no difference in side-effect profile and four studies did not report details of side-effects.

### Sub-group 4: Subcutaneous adrenaline compared to parenteral selective β_2_-agonist

Thirteen studies compared subcutaneous adrenaline and parenteral (subcutaneous or intravenous) selective β_2_-agonist. All 13 reported similar initial bronchodilation with adrenaline or selective β_2_-agonist although one study noted that by 90 minutes the bronchodilation caused by adrenaline was significantly less than that caused by salbutamol. Nine studies reported no difference in side effects, two reported greater side-effects (agitation, “excitement”, and headache) with adrenaline, and one reported mild tremor in two patients in the salbutamol group.

### Sub-group 5: Subcutaneous adrenaline compared to oral selective β_2_-agonist

One study (Calvo et al. 1982) compared subcutaneous adrenaline and oral fenoterol and reported greater bronchodilation and a favourable side-effect profile with fenoterol.

### Sub-group 6: Nebulised adrenaline compared to nebulised selective β_2_-agonist

Six studies explored this comparison and all showed equivalent efficacy. Three studies reported equivalent side-effect profile between groups, two reported increased frequency of mild side-effects with adrenaline and one did not report data on side-effects.

### Unpublished studies

A search of the WHO International Clinical Trials Registry Platform identified one unpublished study of intramuscular adrenaline in addition to selective β_2_-agonist in 49 children aged 6 to 18 with a severe asthma exacerbation, NCT01705964. This study has been completed but not yet been reported and results are not available on the clinicaltrials.gov registry.

## Discussion

This systematic review and meta-analysis of randomised controlled trials of adrenaline versus selective β_2_-agonist for acute asthma found no evidence of a different risk of treatment failure for adrenaline by any route compared to administration of selective β_2_-agonist by any route. However, the individual studies were largely at high risk of bias, there was evidence of statistical heterogeneity, and the studies were clinically heterogeneous. Meta-regression provided weak evidence that some of the heterogeneity was explained by the route of administration of intervention and control, with parenteral routes of administration associated with greater risk of treatment failure. There was strong evidence that the recruitment age-group was associated with different estimates of the risk of treatment failure; with the studies recruiting adults-only having a lower risk of treatment failure with adrenaline. It is unclear whether this represents a genuine difference in treatment response between adults and children or simply reflects underlying differences in the design, setting, and study methodology. Reporting of important clinical outcomes, here considered secondary outcomes, was very poor and precluded a pooled analysis, however a narrative review was consistent with no difference in other outcomes.

It was not possible to determine whether the addition of intramuscular adrenaline to nebulised selective β_2_-agonist improves outcomes in patients with acute asthma. There is very limited evidence from two unblinded studies of the addition of subcutaneous adrenaline to nebulised selective β_2_-agonist, which suggests that the addition of subcutaneous adrenaline did not reduce treatment failure or lead to greater bronchodilation than nebulised selective β_2_-agonist alone. The established pharmacokinetics of adrenaline versus other selective β_2_-agonists means that adrenaline dosing is required more frequently, typically every 20 minutes.[14]

There is inconsistent and low-quality evidence that parenteral adrenaline has a worse side-effect profile than parenteral selective β_2_-agonists. Side-effect data was inconsistently reported however agitation, tremor and headache appeared more frequently with adrenaline. Tachycardia was not consistently reported to be present more frequently with adrenaline and the data was insufficient to assess whether cardiovascular side-effects were more common with adrenaline.

It is appropriate to highlight that adrenaline was the reference treatment at the time many of the studies included in this review were published, with selective β_2_-agonist less frequently used, and so the purpose of the majority of studies was to compare the efficacy and side-effect profile of selective β_2_-agonists to that standard. This may explain why the clinical scenario of most relevance today, addition of intramuscular adrenaline to selective β_2_-agonist, was rarely addressed.

We did not limit our search by language and location although we were only able to include studies written in English, French or Spanish, and so excluded 15 studies. A limitation of this review is that the overall low quality of evidence with marked clinical heterogeneity precluded meta-analysis of secondary outcome data. A weakness of this analysis is that some of the sub-groups only had one or two studies contributing data and so estimates of heterogeneity explicable by these sub-groups and the effects within sub-groups may not be reliable. The findings of this review are limited to the doses and routes studied in the included trials. Due to the limited evidence base, we are unable to draw definitive conclusions on the efficacy of adrenaline in comparison to selective β_2_-agonist in the treatment of acute asthma exacerbations and there is clinical equipoise as to whether addition of adrenaline to selective β_2_-agonist improves outcome compared to selective β_2_-agonist alone.

There is a clear difference between the management recommended in national and international asthma guidelines, where adrenaline use is only recommended if there are features of anaphylaxis or angioedema, and that recommended in many pre-hospital care ambulance guidelines, where intramuscular adrenaline is recommended in severe or life-threatening asthma. There is therefore an urgent need for high quality evidence to determine whether there is a role for adrenaline in the management of severe and life-threatening asthma. If there is a benefit, asthma guidelines and clinical practice in hospital will need to be updated to include the use of parenteral adrenaline in addition to selective β_2_-agonist in this circumstance. Conversely, if there is no evidence of benefit updates to pre-hospital guidelines will be necessary to advise against the use of parenteral adrenaline in the absence of anaphylaxis.

### Conclusions

The limited evidence available suggests that adrenaline and selective β_2_-agonists have similar efficacy in acute asthma but that adrenaline has a worse side-effect profile. There was no evidence of benefit from the use of adrenaline in addition to selective β_2_-agonists in acute asthma but the available evidence is insufficient to rule out a clinically important benefit. There is clinical equipoise as to whether addition of adrenaline improves outcomes and a need for high-quality double-blind RCTs to address this issue. The primary question that studies should aim to address is whether the addition of intramuscular adrenaline to inhaled selective β_2_-agonist reduces the risk of treatment failure or death in adults or children with acute asthma.

## Data Availability

The data that support the findings of this study are available from the corresponding author upon reasonable request

## Acknowledgements

The authors would like to thank Denise Fabian and Jung Cho for assistance obtaining journal articles, Gwendoline Peter for assistance translating one of the manuscripts and Andrew Davies for clinical conversations which helped conceive this research question.

## Contribution

JF, CB, DS, JH and RB proposed the systematic review. JF, CB, DS, JH and JS screened the studies and extracted the data. MW conducted the meta-analysis. JF wrote the first draft of the manuscript. All authors had access to the data and read and approved the final version of the manuscript. JF is guarantor of the data.

## References

1 Arthur G. Epinephrine: a short history. 2015. doi:10.1016/S2213-2600(15)00087-9

2 Melland B. The treatment of spasmodic asthma by the hypodermic injection of adrenalin. Lancet 1910;175:1407–11. doi:10.1016/S0140-6736(01)14446-6

3 Holley AD, Boots RJ. Review article: Management of acute severe and near-fatal asthma. Emerg Med Australas 2009;21:259–68. doi:10.1111/j.1742-6723.2009.01195.x

4 Long B, Lentz S, Koyfman A, et al. Evaluation and management of the critically ill adult asthmatic in the emergency department setting. Am J Emerg Med Published Online First: 19 March 2020. doi:10.1016/j.ajem.2020.03.029

5 National Clinical Guideline Centre. Asthma: diagnosis and monitoring of asthma in adults, children and young people. https://www.nice.org.uk/guidance/gid-ng10095/ (accessed 10 Nov 2017).

6 Beasley R, Beckert L, Fingleton J, et al. Asthma and Respiratory Foundation NZ Adolescent and Adult Asthma Guidelines 2020: a quick reference guide. NZMJ 2020;26:1517.www.nzma.org.nz/journal (accessed 23 Oct 2020).

7 National Asthma Council Australia. Australian Asthma Handbook, Version 2.1. Melbourne: 2020. http://www.asthmahandbook.org.au

8 National Heart Lung and Blood Institute (NHLBI). Expert Panel Report 3: Guidelines for the Diagnosis and Management of Asthma Full Report 2007. Published Online First: 2007.https://www.nhlbi.nih.gov/sites/default/files/media/docs/asthgdln_1.pdf

9 Wellington Free Ambulance. Wellington Free Ambulance Clinical Procedures and Guidelines. 2019.

10 St John New Zealand. Clinical Procedures and Guidelines. 2019.

11 Joint Royal Colleges Ambulance Liason Committee (JRCALC), Association of Ambulance Chief Executives (AACE). JRCALC Clinical Guidelines 2019. 2019. https://aace.org.uk/clinical-practice-guidelines/ (accessed 23 Oct 2020).

12 NASEMSO Medical Directors Council. National Model EMS Clinical Guidelines. 2017. www.nasemso.org (accessed 23 Oct 2020).

13 Secombe P, Stewart P, Singh S, et al. Clinical management practices of life-threatening asthma?: an audit of practices in intensive care. 2019;21.

14 Sellers WFS. Inhaled and intravenous treatment in acute severe and life-threatening asthma. doi:10.1093/bja/aes444

15 Nakamura, Y., Tamaoki, J., Nagase, H., Yamaguchi, M., Horiguchi, T., Hozawa, S., Ichinose, M., Iwanaga, T., Kondo, R., Nagata, M., Yokoyama, A., & Tohda, Y. (2020). Japanese guidelines for adult asthma 2020. Allergology International, 69(4), 519–548. https://doi.org/https://doi.org/10.1016/j.alit.2020.08.001

16 Stanley D, Frca M, Tunnicliffe W. Management of life-threatening asthma in adults. doi:10.1093/bjaceaccp/mkn012

17 Arrest Asthma - RCEMLearning. https://www.rcemlearning.co.uk/foamed/arrest-asthma/ (accessed 23 Oct 2020).

18 Fingleton J, Baggott C, Sabbagh D, et al. Protocol for a systematic review of the safety and efficacy of adrenaline therapy administered by any route for acute severe exacerbations of asthma. 2017. https://www.crd.york.ac.uk/PROSPERO/display_record.php?ID=CRD42017079472

19 Moher D, Shamseer L, Clarke M, et al. Preferred reporting items for systematic review and meta-analysis protocols (PRISMA-P) 2015 statement. Syst Rev 2015;4:1.https://systematicreviewsjournal.biomedcentral.com/articles/10.1186/2046-4053-4-1 (accessed 23 May 2017).

20 Veritas Health Innovation. Covidence systematic review software. www.covidence.org

21 Higgins JPT, Altman DG, Gøtzsche PC, et al. The Cochrane Collaboration’s tool for assessing risk of bias in randomised trials. BMJ 2011;343. doi:10.1136/bmj.d592822

22 Kornberg, A., Zuckerman, SJRW., Mezzadri, F., & Aquino, N. (1991). Effect of injected long-acting epinephrine in addition to aerosolized albuterol in the treatment of acute asthma in children. Pediatric Emergency Care, 7(1 PG-1–3), 1–3.

23 Quadrel M, Lavery RF, Jaker M, et al. Prospective, randomized trial of epinephrine, metaproterenol, and both in the prehospital treatment of asthma in the adult patient. Ann Emerg Med 1995;26:469– 473.http://www.sciencedirect.com/science/article/pii/S0196064495701168 (accessed 22 May 2017).

24 Turpeinen M, Kuokkanen J, Backman A. Adrenaline and nebulized salbutamol in acute asthma. Arch Dis Child 1984;59:666–8. doi:10.1136/adc.59.7.666

25 Naspitz C, Solé D, Wandalsen N. Treatment of acute attacks of bronchial asthma. A comparative study of epinephrine (subcutaneous) and fenoterol (inhalation). Ann Allergy 1987;59:21–4.

26 Uden D, Goetz D, Kohen D, et al. Comparison of nebulized terbutaline and subcutaneous epinephrine in the treatment of acute asthma. Ann Emerg Med 1985;14:229–32.

27 Lin Y, Kh H, Lf C, et al. Terbutaline nebulization and epinephrine injection in treating acute asthmatic children. Pediatr allergy Immunol 1996;7:95–9.

28 Becker, A. B., & Simons, E. R. (1962). Inhaled salbutamol (Albuterol) vs injected epinephrine in the treatment of acute asthma in children. Journal of Allergy and Clinical Immunology, 69(1), 136. https://doi.org/10.1016/S0091-6749(62)80456-4

29 Ruddy, R., Kolski, G., Scarpa, N., & Wilmott, R. (1986). Aerosolized metaproterenol compared to subcutaneous epinephrine in the emergency treatment of acute childhood asthma. Pediatric Pulmonology, 2(4 PG-230–236), 230–236.

30 Ibrahim, S. A., Elgurashi, E. D., & Elkarim, O. A. (1993). Comparative study of intravenous aminophylline subcutaneous adrenaline and nebulised salbutamol in the treatment of acute asthma in children. Pediatric Reviews and Communications, 7(3), 175–182.

31 Sharma A, Madan A. Subcutaneous epinephrine vs nebulized salbutamol in asthma. Indian J Pediatr 2001;68:1127–30.

32 Ben-Zvi Z, Lam C, Hoffman J, et al. An evaluation of the initial treatment of acute asthma. Pediatrics 1982;70:348–53.

33 Pancorbo S, Fifield G, Davies S, et al. Subcutaneous epinephrine versus nebulized terbutaline in the emergency treatment of asthma. Clin Pharm 1983;2:45–8.

34 Tinkelman DG, Vanderpool GE, Carroll MS, et al. Comparison of nebulized terbutaline (TERB) and subcutaneous epinephrine (EPI) in the treatment of acute asthma. Ann Allergy 1983;50:398–401.

35 Phanichyakarn P, Ananthachai C, Direkwattanachai C. Adrenaline and terbutaline in treatment of acute asthmatic attacks in children. J Med Assoc Thail 1981;64:428–31.

36 Elenbaas RM, Frost GL, Robinson WA, et al. Subcutaneous Epinephrine Vs. Nebulized Metaproterenol in Acute Asthma. Drug Intelligence & Clinical Pharmacy. 1985;19(7-8):567–571. doi:10.1177/106002808501900714

37 Schwartz AL, Lipton JM, Warburton D, et al. Management of Acute Asthma in Childhood: A Randomized Evaluation of β-Adrenergic Agents. Am J Dis Child 1980;134:474–8. doi:10.1001/archpedi.1980.02130170024009

38 Appel D, Jp K, Sherman M. Epinephrine improves expiratory flow rates in patients with asthma who do not respond to inhaled metaproterenol sulfate. J Allergy Clin Immunol 1989;84:90–8.

39 Youngchaiyud P, Charoenratanakul S. Terbutaline pressurised aerosol inhaled via a Nebuhaler--an effective alternative to subcutaneous adrenaline for treatment of acute severe asthma. Eur J Respir Dis 1987;70:284–92.

40 Lopez-Messa JB, Reyes A, Alonso P, et al. Subcutaneous epinephrine (adrenaline) vs intravenous salbutamol in acute severe asthma. Clin Intensive Care 1992;3:153–9.

41 Giner MT, Nevot S, Sierra JI, Plaza A, Coma G, Youssef W. Tratamiento de las crisis agudas de asma bronquial en niños con salbutamol subcutáneo como alternativa a la adrenalina: estudio comparativo [Treatment of acute crises of bronchial asthma in children using subcutaneous salbutamol as an alternative to adrenaline: comparative study]. An Esp Pediatr. 1986 Sep;25(3):165–9. Spanish.

42 Phanichyakarn P. Comparison of subcutaneous injections of tubutaline, salbutamol and adrenaline in acute asthmatic attacks in children. J Med Assoc Thail 1989;72:692–6.

43 Khaldi F, Salem N. Comparison of the effect of subcutaneous injection of adrenaline and terbutaline in asthma crisis in infants. Arch Pediatr 1998;5:745–8.

44 Sly R, Badiei B, Faciane J. Comparison of subcutaneous terbutaline with epinephrine in the treatment of asthma in children. J Allergy Clin Immunol 1977;59:128–35.

45 Davis W, Pang L, Chernack W, et al. Terbutaline in the treatment of acute asthma in childhood. Chest 1977;72:614–7.

46 Simons F, Gillies J. Dose response of subcutaneous terbutaline and epinephrine in children with acute asthma. Am J Dis Child 1981;135:214–7.

47 Ferres J, Ma M, Farre R, et al. Subcutaneous epinephrine versus inhaled salbutamol in the treatment of acute asthma in pediatrics. An españoles pediatría 1987;27:37–40.

48 Smith P, Heurich A, Leffler C, et al. A comparative study of subcutaneously administered terbutaline and epinephrine in the treatment of acute bronchial asthma. Chest 1977;71:129–34.

49 Spiteri, M. A., Millar, A. B., Pavia, D., & Clarke, S. W. (1988). Subcutaneous adrenaline versus terbutaline in the treatment of acute severe asthma. Thorax, 43(1), 19–23. https://doi.org/10.1136/thx.43.1.19

50 Aggarwal P, Jn P, Js G. Bronchodilators in acute bronchial asthma?: a comparative study. Indian J Chest Dis Allied Sci 1986;28:21–7.

51 Baughman R, Ploysongsang Y, James W. A comparative study of aerosolized terbutaline and subcutaneously administered epinephrine in the treatment of acute bronchial asthma. Ann Allergy 1984;53:131–4.

52 Calvo M, Escobar L. Ambulatory treatment of patients with asthmatic crisis: comparison of two therapeutic regimens. Rev Chil Pediatr 1982;53:107–10.

53 Khaldi F, Tabarki B, Salem N. Comparison of inhaled nebulised adrenaline with salbutamol in acute asthma. Saudi Med J 1998;19:409–12.

54 Plint AC, Osmond MH, Klassen TP. The efficacy of nebulized racemic epinephrine in children with acute asthma: A randomized, double-blind trial. Acad Emerg Med 2000;7:1097–103. doi:10.1111/j.1553-2712.2000.tb01258.x

55 Adoun M, Jp F, Doré P, et al. Comparison of nebulized epinephrine and terbutaline in patients with acute severe asthma: a controlled trial. J Crit Care 2004;19:99–102.

56 Abroug F, Nouira S, Bchir A, et al. A controlled trial of nebulized salbutamol and adrenaline in acute severe asthma. Intensive Care Med 1995;21:18–23.

57 Zeggwagh A, Abouqal R, Madani N, et al. Comparative efficiency of nebulized adrenaline and salbutamol in severe acute asthma. A randomized, controlled prospective study. Ann Fr Anesth Reanim 2002;21:703–9.

58 Coupe M, Guly U, Brown E, et al. Nebulished adrenaline in acute severe asthma: comparison with salbutamol. Eur J Respir Dis 1987;71:227–32.

