## Supplementary Appendix for "Adrenaline (epinephrine) compared to selective beta-2-agonist in adults or children with acute asthma: a systematic review and meta-analysis"

### Supplementary appendix: Systematic review of the comparison between adrenaline (epinephrine) and selective beta-2 agonist in the setting of adults or children with acute asthma

**Authors:**

[1]Christina Baggott, [1]Jo Hardy, [1]Jenny Sparks, [1]Doñah Sabbagh, [1,2]Richard Beasley, [3]Mark Weatherall, [1,2,3]James Fingleton

**Author's affiliations**

[1]Medical Research Institute of New Zealand, Wellington, New Zealand

[2]Capital and Coast District Health Board, Wellington, New Zealand

[3]University of Otago Wellington, Wellington, New Zealand

**Word count:**

**Key words:** Asthma, adrenaline, epinephrine, beta-agonists, systematic review, meta-analysis

**Corresponding author**

James Fingleton
Medical Research Institute of New Zealand
Private Bag 7902, Newtown, Wellington 6242, New Zealand
Facsimile: +64-4-389 5707

#### Information Sources and Search Strategy

We searched the Cochrane Central Register of Controlled Trials and Scopus databases. Reference lists from included studies and known reviews were also hand searched. The searches were originally conducted 15th June 2018 and a repeat search on 23rd October 2020 identified no additional relevant studies.

**Scopus search terms:**
( TITLE-ABS-KEY ( asthma OR asthmatic ) AND TITLE-ABS-KEY ( adrenaline OR epinephrine ) ) AND ( LIMIT-TO ( DOCTYPE , "ar" ) ) AND ( LIMIT-TO ( EXACTKEYWORD , "Human" ) )

**Cochrane Central Register of Controlled Trials Airways Group Register search terms:**

(asthma OR asthmatic) AND (adrenaline OR epinephrine):ti,ab,kw" in Trials (Word variations have been searched)

**Table S1: Summary of treatment failure and lung function data for all included studies**

| Study | Treatment failure summary | Lung function summary | Summary of authors overall conclusion |
|---|---|---|---|
| **Subgroup 1: Subcutaneous adrenaline in addition to inhaled/nebulised selective β2 agonist versus inhaled/nebulised selective β2 agonist** | | | |
| Kornberg, 1991 | 1/20 patient who received long acting adrenaline and 3/23 patients who received placebo in addition to salbutamol were admitted. | No significant difference in PEF between group that received long acting adrenaline or placebo in addition to nebulised salbutamol | Long acting adrenaline provided no additional benefit to nebulised salbutamol |
| Quadrel, 1995 | There was no significant difference in hospitalisation between the treatment groups. Two patients were intubated; 1/53 in the adrenaline group and 1/49 in the metaproterenol group | Peak flow did not differ significantly between treatment groups | There was no significant difference in outcome between the groups. |
| **Subgroup 2: Intramuscular adrenaline versus inhaled/nebulised selective β2 agonist** | | | |
| Turpeinen, 1984 | 9/20 patients in the adrenaline group and 5/26 in the salbutamol group were admitted to hospital | PEFR improved significantly in both groups, however improvements were significantly greater with salbutamol than adrenaline. | Both injected adrenaline and inhaled salbutamol have good bronchodilatory effect. Salbutamol was significantly more effective than adrenaline. |
| **Subgroup 3: Subcutaneous adrenaline versus inhaled/nebulised selective β2 agonist** | | | |
| Naspitz, 1984 | No significant difference in number of participants requiring additional treatment (32/75 in adrenaline group versus 22/75 in Fenoterol group). | No significant difference in clinical score between the two groups | Adrenaline and fenoterol were equally effective |
| Uden, 1985 | 1/12 participants in adrenaline group and 0/7 in terbutaline group were admitted | FEV1 was not significantly different between the two groups | Nebulised terbutaline was as effective as sc adrenaline |
| Lin, 1996 | Not reported | FEV1 was significantly improved in all treatment groups | Both treatments were effective bronchodilators. Adrenaline was more effective but with greater side-effects. Authors recommended β2 agonist as first line therapy. |
| Becker, 1983 | No significant difference between groups with regard to repeated treatment or admission, numbers not reported. | Significant improvement in FEV1 with no significant difference between groups | Inhaled nebulized salbutamol and subcutaneous epinephrine are equally effective. Salbutamol had fewer side-effects |
| Ruddy, 1986 | 1/19 patient in the metaprotenerol group and 4/16 in the adrenaline group required further therapy or hospitalisation | Significant improvement in PEFR & FEV1 with no significant difference between groups | Metaproterenol was as good as adrenaline with few side effects in both groups |
| Ibrahim, 1993 | Not reported | Significant improvement in PEFR in all groups, PEFR was significantly higher in the salbutamol group at 10 and 30 minutes | Treatment with salbutamol is superior to adrenaline with fewer side effects |
| Sharma, 2001 | 4/25 patients in each treatment groups failed to respond to treatment | Increase in PEFR was similar between all treatment groups | Adrenaline and salbutamol resulted in equal improvement in PEFR |

| | | | |
|---|---|---|---|
| Ben-Zvi, 1982 | 4/24 patients in the adrenaline group and 0/26 patients in the fenoterol group had treatment failure or required further treatment. | There was greater improvement in FEV1 with fenoterol than adrenaline | Adrenaline was associated with significantly more treatment failure and side effects |
| Pancorbo, 1983 | 4/11 patients in the adrenaline group and 5/10 in the terbutaline group had poor improvement in PEFR | There was a significantly greater improvement in PEF at 5 minutes with terbutaline than adrenaline however there was no significant difference at 15 or 30 minutes | Terbutaline is as effective as adrenaline |
| Tinkelman, 1983 | 1/15 patients in the terbutaline group and 0/18 in the adrenaline group did not have a significant clinical improvement | Significant improvement in FEV1 in both treatment groups with no significant difference between them | Both treatments were equally effective with no significant difference in effectiveness or side effects |
| Anantharaman, 1993 | 4/27 of patients in the adrenaline group and 6/17 in the salbutamol group required hospitalisation. | Greater in PEFR improvement with salbutamol than adrenaline, however this was not statistically significant | Salbutamol and adrenaline are effective for treating acute asthma |
| Elenbaas, 1985 | Not reported | Both treatment groups had a significant increase in % predicted PEFR with no difference between groups | Adrenaline and metaproterenol appear to be equally effective |
| Schwartz, 1980 | 15/66 patients in the adrenaline group and 34/127 in the neb isoetharine group were admitted | Significant increase in FEV1 with no significant difference between the groups | All beta adrenergic agents were equally effective |
| Appel, 1989 | 6/54 patients in the adrenaline group and 18/46 in the metaproterenol group failed to improve | PEFR improved more rapidly in patients treated initially with adrenaline than metaproterenol | Adrenaline was often the more effective bronchodilator |
| Youngchaiyud, 1987 | 1 patient from the adrenaline group was excluded from the analysis for failing to respond and being unable to perform lung function testing. No other data. | Greater improvement in FEV1 with terbutaline than adrenaline, this was not statistically significant | Terbutaline was as effective as adrenaline with fewer side effects |
| Ferres, 1987 | Not reported | Decrease in asthma severity score in all groups, with no significant difference between groups | Salbutamol was as effective as adrenaline with fewer side effects |
| **Subgroup 4: Subcutaneous adrenaline versus intravenous/subcutaneous selective β2 agonist** | | | |
| Lopez-Messa, 1992 | Not reported | No significant difference in FEV1 or PEFR between both treatment groups | Adrenaline and salbutamol had similar efficacy with few and mild side effects |
| Giner, 1986 | 3/15 patients in the adrenaline group and 5/15 in the salbutamol group were admitted for progressive deterioration | Statistically significant increase in FEV1 in both groups, increase was maintained to 90mins in the salbutamol group but not in the adrenaline group | Sc salbutamol is preferred to sc adrenaline |
| Phanichyakarn, 1989 | Not reported | Significant increase in PEF for all groups. Greatest in the terbutaline group but this was not statistically significant | All drugs were effective but terbutaline led to greater bronchodilation than salbutamol or adrenaline |
| Khaldi, 1998 | Not reported | Lung function tests not undertaken. Reported improvement in clinical signs and symptoms, oxygen saturations and PaO2 were similar between both groups. | Adrenaline was as effective as terbutaline in treating acute asthma |

| | | | |
|---|---|---|---|
| Sly, 1977 | 5/20 patients in adrenaline group and 1/20 patient in terbutaline group did not respond | Significant increase in PEF with no significant difference between the two groups | Both treatments were equally effective |
| Davis, 1977 | Not reported | Greater improvement in FEV1 with terbutaline than adrenaline but limited data | Both treatments were equally effective but adrenaline had greater side-effects |
| Simons, 1981 | No patient was withdrawn for increasing respiratory distress | Significant increase in FEV1 with no significant difference between the four groups | Terbutaline and adrenaline were equally effective but adrenaline had more side-effects |
| Phanichtakarn, 1981 | Not reported | Significant increase in PEF with no significant difference between the two groups | No significant difference in efficacy between the two groups but more cardiovascular side effects were observed with adrenaline. |
| Smith, 1977 | Not reported | Significant improvement in FEV1 with no significant difference between two treatment groups | Terbutaline is an effective bronchodilator |
| Spiteri, 1988 | Not reported | Increase in PEF and FEV1 with no significant between treatment groups | Adrenaline and terbutaline lead to rapid effective bronchodilation without important side effects |
| Aggarwal, 1986 | Not reported | Significant improvement in PEF not significantly different between groups | Adrenaline, salbutamol and terbutaline were equally effective |
| Baughman, 1984 | Not reported | Significant improvement in FEV1 and PEFR in both groups, with no significant difference between treatments | Terbutaline has similar effectiveness to adrenaline |
| **Subgroup 5: Subcutaneous adrenaline versus oral selective β2 agonist** | | | |
| Calvo, 1982 | Not reported | Siginficant increase in Maximum Expiratory Flow in both groups with a significantly greater increase with fenoterol | Fenoterol was superior to adrenaline for bronchodilation and side-effect profile. |
| **Subgroup 6: Nebulised adrenaline versus inhaled/nebulised selective β2 agonist** | | | |
| Khaldi, (2) 1998 | Not reported | All patients had an increase in peak with no difference between the two groups. | Nebulised adrenaline was as effective as a nebulised beta agonist in acute asthma without significant cardiovascular side effects. |
| Plint, 2000 | 8/61 patients from the salbutamol group and 9/60 from the adrenaline group were hospitalised | No significant difference between the two groups in pulmonary severity index at any time point. | Equal benefits of both drugs |
| Adoun, 2004 | 4/18 patients not improved in the adrenaline group, 9/20 patients not improved in the salbutamol group | PEF improved significantly in both groups | No synergistic effect from adrenaline following terbutaline. Both adrenaline and terbutaline were equally effective and well tolerated |
| Abroug, 1995 | No patients (0/11 in each group) required intubation or died | PEF improved in both groups with no significant differences between the groups | Nebulised adrenaline was as safe and effective as nebulised salbutamol |
| Zeggwagh, 2002 | No patients were intubated (0/22 per group) | Significant increase in PEF with no significant difference between the two groups | Nebulised adrenaline was as effective as nebulised salbutamol without significant side effects |
| Coupe, 1987 | Not reported | PEF increased in all patients with no significant difference between the two groups | Nebulised adrenaline was an effective bronchodilator but did not produce a significantly greater bronchodilatory response than salbutamol |

**Table S2: Heart rate and adverse events**

| Study | Heart rate summary | Adverse event summary | Summary of authors overall conclusion |
|---|---|---|---|
| **Subgroup 1: Subcutaneous adrenaline in addition to inhaled/nebulised selective β2 agonist versus inhaled/nebulised selective β2 agonist** | | | |
| Kornberg, 1991 | Heart rate declined in both groups | Not mentioned in text | Not mentioned in text |
| Quadrel, 1995 | Heart rate did not differ significantly between the groups | Not mentioned in text | No side effects were observed in the treatment groups |
| **Subgroup 2: Intramuscular adrenaline versus inhaled/nebulised selective β2 agonist** | | | |
| Turpeinen, 1984 | No significant change in heart rate in either group | Not mentioned in text | Muscle tremor in 5 patients in the adrenaline group and 2 in the salbutamol group |
| **Subgroup 3: Subcutaneous adrenaline versus inhaled/nebulised selective β2 agonist** | | | |
| Naspitz, 1984 | Similar in both groups | Not mentioned in text | Not mentioned in text |
| Uden, 1985 | Similar in both groups | Not mentioned in text | Not mentioned in text |
| Lin, 1996 | Similar in both groups | Not mentioned in text | Adverse events in 21/45 those given adrenaline and 5/45 of those given terbutaline. This was statistically significant. |
| Becker, 1983 | Significant increase in heart rate with salbutamol but not with adrenaline | Sinus tachycardia with salbutamol | "Adverse effects, including nausea, vomiting, tremor, headache, palpitations, excitement, and pallor, were seen in 10 of 20 patients given epinephrine. No side effects were seen in those given salbutamol" |
| Ruddy, 1986 | Heart rate was similar in both groups and unchanged from baseline | 1 participant in metaproterenol group had palpitations | None had major side effects necessitating discontinuation of the study |
| Ibrahim, 1993 | Significantly more tachycardia in the adrenaline group | Palpitations occurred exclusively in the adrenaline group | Tremors in 6 patients in the salbutamol group and 21 in the adrenaline group |
| Sharma, 2001 | Heart rate was significantly higher at 30 minutes but not at other times points with adrenaline | 13 patients in the adrenaline group and 14 patients in the terbutaline group had palpitations | 6 patients in adrenaline group and 8 patients in terbutaline group had tremors |
| Ben-Zvi, 1982 | Heart rate did not differ significantly between the groups | Not mentioned in text | 8 patients in adrenaline group and 5 patients in fenoterol group experienced side effects |
| Pancorbo, 1983 | Heart rate did not differ significantly between the groups | Not mentioned in text | Not mentioned in text |
| Tinkelman, 1983 | No significant change from baseline in either group | Not mentioned in text | Side effects were similar between groups with no significant differences between groups |
| Anantharaman, 1993 | Heart rate was similar in both treatment groups | No cardiovascular side effects were observed | Not mentioned in text |
| Elenbaas, 1985 | Heart rate was unchanged in the adrenaline group but decreased significantly in the metaproterenol group | Not mentioned in text | No significant difference in frequency of side effects between the two treatment groups |
| Schwartz, 1980 | Increase in heart rate more frequent with terbutaline | Not mentioned in text | Higher frequency of mild side effects such as tremor and increased heart rate with terbutaline than adrenaline or isoproterenol |

| | | | |
|---|---|---|---|
| Appel, 1989 | Heart rate was similar in both treatment groups | Not mentioned in text | No major side effects were observed |
| Youngchaiyud, 1987 | Significant decrease in heart rate in both groups | Not mentioned in text | Incidence of tremor with adrenaline was twice that of terbutaline, incidence of other side effects was low |
| Ferres, 1987 | Significantly more tachycardia with adrenaline | Not mentioned in text | No important side effects were observed in any of the three groups |
| **Subgroup 4: Subcutaneous adrenaline versus intravenous/subcutaneous selective β2 agonist** | | | |
| Lopez-Messa, 1992 | Small significant increase in heart rate with salbutamol, similar increase with adrenaline but was not significant. No significant differences between groups | One patient in each treatment group had occasional supraventricular beats | Side effects were mild and infrequent in both groups |
| Giner, 1986 | With salbutamol small significant increase in HR, HR increase with adrenaline was not significant | Not mentioned in text | Mild tremor in 2 patients in the salbutamol group |
| Phanichyakarn, 1989 | Significantly more tachycardia in the salbutamol group | Not mentioned in text | No major side effects noted |
| Khaldi, 1998 | Similar in both groups | Not mentioned in text | No adverse events in either group |
| Sly, 1977 | Small significant increase in HR with terbutaline but not with adrenaline | Not mentioned in text | No clinically significant side effects in either group |
| Davis, 1977 | Heart rate increased with terbutaline but there was no significant difference between groups | Not mentioned in text | Mild side effects in 4 patients receiving terbutaline and 5 receiving adrenaline |
| Simons, 1981 | Increased significantly with time in all groups | No patients complained of palipiatations | “Excitement” and headache in 50% of patients receiving adrenaline and not with terbutaline. Agitation with an unspecified number of patients given adrenaline. Tremor in patients given adrenaline and highest doses of terbutaline |
| Phanichtakarn, 1981 | Increased in both groups, numerically greater increase with adrenaline | Increased heart rate and blood pressure with adrenaline but no serious side effects reported | No severe side effects noted |
| Smith, 1977 | Heart rate in terbutaline group was greater than with the adrenaline group | Not mentioned in text | Equal frequency of side effects between the two treatment groups |
| Spiteri, 1988 | No significant increase in heart rate with either treatment | Not mentioned in text | No important side effects |
| Aggarwal, 1986 | Decline in heart rate in all treatment groups | Not mentioned in text | Minor side effects not severe enough to necessitate termination |
| Baughman, 1984 | Heart rate was significantly lower with terbutaline | Not mentioned in text | 3 patients in adrenaline group and 0 patients in the terbutaline group had mild side effects |
| **Subgroup 5: Subcutaneous adrenaline versus oral selective β2 agonist** | | | |
| Calvo, 1982 | Decreased HR in both groups | Not mentioned in text | Fenoterol was better tolerated with fewer side effects |
| **Subgroup 6: Nebulised adrenaline versus inhaled/nebulised selective β2 agonist** | | | |
| Khaldi, (2) 1998 | Similar in both groups | No significant cardiovascular side effects in either group | Not mentioned in text |
| Plint, 2000 | No significant difference in heart rate | Not mentioned in text | Significantly higher incidence of nasal pharyngeal side effects with adrenaline. |

| | | | |
|---|---|---|---|
| Adoun, 2004 | Similar in both groups | Not mentioned in text | Both treatments were well tolerated |
| Abroug, 1995 | Similar in both groups | Arrhythmias and tachycardia occurred with equal frequency in both groups. | Side effects were mild and slightly more frequent with adrenaline |
| Zeggwagh, 2002 | Heart rate slowed in both groups | Not mentioned in text | No significant difference in side effects between the two groups |
| Coupe, 1987 | Similar in both groups | Not mentioned in text | No significant difference in side effects between the two groups |

**Table S3: Table of excluded studies**

| Study | Reason for exclusion |
|---|---|
| Ablakulova 1963 | Not in a language we can translate |
| Abramson 1963 | Not an RCT |
| Abroug 2016 | Not an RCT |
| Adam 1938 | Not an RCT |
| Ahmad 1980 | Not an RCT |
| Alliott 1972 | Not acute asthma |
| Andre 1951 | Not acute asthma |
| Appel 1978 | Conference paper/abstract |
| Appel 1981 | No selective beta 2 agonist comparator |
| Bacq 1961 | Not an RCT |
| Badatcheff 1989 | Conference paper/abstract |
| Barlow 1930 | Not an RCT |
| Becker 1982 | Conference paper/abstract |
| Ben Zvi 1981 | Conference paper/abstract |
| Ben Zvi 1983 | No selective beta 2 agonist comparator |
| Bleecker 1969 | Wrong comparator |
| Bookman 1980 | Not an RCT |
| Brandon 1977 | Not an RCT |
| Brandstetter 1980 | No selective beta 2 agonist comparator |
| Brogden 1973 | Not an RCT |
| Burton 1982 | Not an RCT |
| Collins Williams 1968 | Not an RCT |
| Costa 1973 | Not acute asthma |
| Coupe 1987a | Duplicate study |
| CoupeM 1986 | Conference paper/abstract |
| Crompton 1968 | No selective beta 2 agonist comparator |
| Cydulka 1988 | No selective beta 2 agonist comparator |
| Dockar 1940 | Not an RCT |
| Douglas 1966 | Not acute asthma |
| Elatrous 1997 | Not acute asthma |
| Elatrous 1997a | Duplicate study |
| EspinosaAyala 1980 | Not acute asthma |
| Fanta 1986 | No selective beta 2 agonist comparator |
| Fireman 1975 | Not an RCT |
| Fossati 1975 | Not an RCT |
| Fourton 1950 | Unable to obtain through inter-library loan |
| Fradkin 1979 | Not in a language we can translate |
| Franchini 2010 | Not an RCT |
| Francis 1933 | Not an RCT |
| Frouchtman 1950 | No selective beta 2 agonist comparator |
| Geisler 1983 | Not in a language we can translate |
| Glass 1973 | Not acute asthma |
| Gotz 1981 | No selective beta 2 agonist comparator |
| Graeser 1935 | No selective beta 2 agonist comparator |
| Grater 1958 | Duplicate study |
| Graterw 1958 | No selective beta 2 agonist comparator |
| Grauvilarrubias 1951 | No selective beta 2 agonist comparator |
| Grebe 1983 | Not in a language we can translate |
| Green 2003 | Not an RCT |
| Hartman 1945 | No selective beta 2 agonist comparator |
| Haughey 1952 | Not an RCT |
| Helander 1967 | No selective beta 2 agonist comparator |
| Herraizballestero 1953 | Not an RCT |

| | |
|---|---|
| Hioco 1951 | Not an RCT |
| Hughes 1978 | Not an RCT |
| Hurst 1940 | Not an RCT |
| Johansen 1974 | Not acute asthma |
| John 2010 | Not acute asthma |
| Johnston 1963 | Not an RCT |
| Josephson 1981 | Not an RCT |
| Kanter 1970 | Not in a language we can translate |
| Karetzky 1974 | No selective beta 2 agonist comparator |
| Karetzky 1978 | No selective beta 2 agonist comparator |
| Karetzky 1980 | No selective beta 2 agonist comparator |
| Kennedy 1964 | No selective beta 2 agonist comparator |
| Kennedy 1972 | Not acute asthma |
| Khaldi 1998b | Duplicate study |
| Khaldi 1998c | Duplicate study |
| Kim 2012 | Not an RCT |
| Kjellman 1980 | Not acute asthma |
| Kobkitsumongkol 1994 | Conference paper/abstract |
| Konig 1978 | Not an RCT |
| Kornberg 1992 | Unable to obtain through inter-library loan |
| Krasner 1940 | Not an RCT |
| Krasner 1941 | Not an RCT |
| Levy 1948 | Not an RCT |
| Lewis 1985 | Not an RCT |
| Limthongkul 1989 | Not an RCT |
| Lindholm 1966 | Not acute asthma |
| Lockey 1943 | No selective beta 2 agonist comparator |
| Lockey 1945 | No selective beta 2 agonist comparator |
| Lopezbarrantes 1951 | Not an RCT |
| Mondal 2014 | Not acute asthma |
| Morice 1986 | Not an RCT |
| Naspitz 1985 | Not in a language we can translate |
| Naspitz 1989 | Not in a language we can translate |
| NCT01705964 2012 | Not an RCT |
| Ngamphaiboon 1989 | Not in a language we can translate |
| Nielsen 1936 | Not an RCT |
| Noseda 1989 | Not an RCT |
| Nu 1971 | Not acute asthma |
| Nutbeam 2009 | Not an RCT |
| Park 1941 | Not an RCT |
| Pavlakou 2004 | Conference paper/abstract |
| Permpikul 1990 | Not in a language we can translate |
| Phanichyakarn 1981a | Duplicate study |
| Phanichyakarn 1989a | Duplicate study |
| Pinnas 1991 | Not acute asthma |
| Pliss 1981 | No selective beta 2 agonist comparator |
| Rees 1967 | Not acute asthma |
| Refsum 1956 | Not in a language we can translate |
| Riding 1970 | Not acute asthma |
| Rohr 1986 | Not acute asthma |
| Ross 1946 | Not an RCT |
| Rossing 1980 | No selective beta 2 agonist comparator |
| Rossing 1981 | No selective beta 2 agonist comparator |
| Royle 1938 | Not an RCT |
| Schwartz 1980a | Duplicate study |
| Sharma 1985 | Not acute asthma |

| | |
|---|---|
| Sly 1969 | No selective beta 2 agonist comparator |
| Snider 1955 | Not acute asthma |
| Streeton 1977 | Not an RCT |
| Taub 1968 | Not an RCT |
| Taub 1971 | Not an RCT |
| Teoh 1979 | Not acute asthma |
| Ting 1991 | Not in a language we can translate |
| Tirot 1992 | Not an RCT |
| Toivonen 1964 | Not in a language we can translate |
| Turner 1938 | Not an RCT |
| Turpeinen 1983 | Not in a language we can translate |
| VonHundelshausen 1983 | Not in a language we can translate |
| Weber 1968 | Not in a language we can translate |
| Weinberger 1974 | No selective beta 2 agonist comparator |
| Zeggwagh 2002a | Duplicate study |

#### References

**Ablakulova 1963**
ABLAKULOVA, Z.; IGNATOVA, Z.G.. COMPARATIVE EVALUATION OF SEVERAL TYPES OF TREATMENT OF PATIENTS WITH BRONCHIAL ASTHMA. Meditsinskii zhurnal Uzbekistana 1963;10:24-26.
**Abramson 1963**
ABRAMSON, H.A.. IMPROVED INHALATION THERAPY OF ASTHMA.. The Journal of asthma research 1963;11:39-48.
**Abroug 2016**
Abroug, F.; Dachraoui, F.; Ouanes-Besbes, L.. Our paper 20 years later: the unfulfilled promises of nebulised adrenaline in acute severe asthma. Intensive Care Medicine 2016;42(3):429-431. [DOI: 10.1007/s00134-016-4210-1]
**Adam 1938**
Adam, J.; Hutchinson, C.A.. Adrenaline Treatment of Asthma. British Medical Journal 1938;2(4067):1279. [DOI: 10.1136/bmj.2.4067.1279-a]
**Ahmad 1980**
Ahmad, M.; Golish, J.A.. Management of difficult asthma. Cleveland Clinic Quarterly 1980;47(1):33-38.
**Alliott 1972**
Alliott RJ; Lang BD; Rawson DR; Leckie WJ. Effects of salbutamol and isoprenaline-phenylephrine in reversible airways obstruction. British medical journal 1972;1(5799):539-542.
**Andre 1951**
ANDRE, M.J.. Sedation of the asthmatic oppression and the dyspnea of the grave cardiac with aerosol-aleudrine inhalation.. Paris médical 1951;41(11):148-152.
**Appel 1978**
Appel, D.; Shim, C.S.; Williams Jr., M.H.. Comparative effect of epinephrine and aminophylline in the treatment of acute asthma. American Review of Respiratory Disease 1978;117(4 II):91.
**Appel 1981**
Appel D; Shim C. Comparative effect of epinephrine and aminophylline in the treatment of asthma. Lung 1981;159(5):243-254.
**Bacq 1961**
BACQ, Z.M.; PHILIPPOT, E.; DEJARDIN, R.; DRESSE, A.. A new interpretation of the resistance to adrenaline in asthmatics.. Revue médicale de Liège 1961;16:497-498.
**Badatcheff 1989**
Badatcheff A; Person C; Raoult J; Meslier N; Racineux JL. Effects of salbutamol and intravenous adrenaline in severe asthmatic crisis. Revue des maladies respiratoires 1989;6(Suppl 3):R168.
**Barlow 1930**
Barlow, O.W.; Frye, J.F.. The antiasthmatic efficiency of epinephrine, ephedrine and atropine: Their comparative effects on a series of experimental attacks in a subject with a complex type of asthma. Archives of Internal Medicine 1930;45(4):538-545. [DOI: 10.1001/archinte.1930.00140100060006]
**Becker 1982**
Becker, A.B.; Estelle, F.; Simons, R.. Inhaled salbutamol (albuterol) vs injected epinephrine in the treatment of acute asthma in children. Journal of Allergy and Clinical Immunology 1982;69(1 II):174.
**Ben Zvi 1981**
Ben-Zvi, Z.; Lam, C.; Spohn, W.. Evaluation of repeated epinephrine injections in acute asthma. American Review of Respiratory Disease 1981;123(4 II):161.
**Ben Zvi 1983**
Ben-Zvi Z; Lam C; Spohn WA; Gribetz I; Mulvihill MN; Kattan M. An evaluation of repeated injections of epinephrine for the initial treatment of acute asthma. American review of respiratory disease 1983;127(1):101-105. [DOI: 10.1164/arrd.1983.127.1.101]

**Bleecker 1969**
Bleecker E; McKinney W; Lyons H; Steen SN. Bronchodilator effects of Quinterenol sulfate--a preliminary clinical study. Anesthesia and analgesia 1969;48(1):7-9.
**Bookman 1980**
Bookman, R.. Epinephrine in acute asthma therapy.. Annals of Allergy 1980;44(4):249.
**Brandon 1977**
Brandon, B.M.; Stolley, P.D.; West, S.K.; Swartz, D.; Rumrill, R.. Drugs used in the treatment of asthma. COMPR.THER. 1977;3(1):9-15.
**Brandstetter 1980**
Brandstetter RD; Gotz VP; Mar DD. Optimal dosing of epinephrine in acute asthma. American journal of hospital pharmacy 1980;37(10):1326-1329.
**Brogden 1973**
Brogden, R.N.; Speight, T.M.; Avery, G.S.. Terbutaline: A Preliminary Report of its Pharmacological Properties and Therapeutic Efficacy in Asthma. Drugs 1973;6(5):324-332. [DOI: 10.2165/00003495-197306050-00002]
**Burton 1982**
Burton, R.M.. Subcutaneous epinephrine for acute asthma. Annals of Emergency Medicine 1982;11(6):335. [DOI: 10.1016/S0196-0644(82)80146-7]
**Collins Williams 1968**
Collins-Williams, C.; Hughes, K.M.. Treatment of the acutely ill asthmatic child.. Annals of Allergy 1968;26(3):117-125.
**Costa 1973**
Costa JL; Goh BK. A comparative trial of subcutaneous terbutaline, Th1165a and adrenaline in bronchial asthma. Medical journal of australia 1973;2(12):588-591.
**Coupe 1987a**
Coupe MO; Guly U; Brown E; Barnes PJ. Nebulised adrenaline in acute severe asthma: comparison with salbutamol. European journal of respiratory diseases 1987;71(4):227-232.
**CoupeM 1986**
Coupe M O; Guly U. Comparison of nebulised adrenaline and salbutamol in acute severe asthma. Clin-sci 1986;71(Suppl 15):80P-81P.
**Crompton 1968**
Crompton, G.K. A comparison of responses to bronchodilator drugs in chronic bronchitis and chronic asthma. Thorax 1968;23(1):46-55. [DOI: 10.1136/thx.23.1.46]
**Cydulka 1988**
Cydulka, R.; Davison, R.; Grammer, L.; Parker, M.; Mathews IV, J.. The use of epinephrine in the treatment of older adult asthmatics. Annals of Emergency Medicine 1988;17(4):322-326. [DOI: 10.1016/S0196-0644(88)80772-8]
**Dockar 1940**
Dockar, A.W.. Adrenaline in Asthma. British Medical Journal 1940;2(4170):807. [DOI: 10.1136/bmj.2.4170.807-a]
**Douglas 1966**
Douglas, A.; Simpson, D.; Merchant, S.; Crompton, G.; Crofton, J.. The effect of antispasmodic drugs on the endomural bronchial (or "squeeze") pressures in bronchitis and asthma.. American Review of Respiratory Disease 1966;93(5):703-715.
**Elatrous 1997**
Elatrous, S.; Elidrissi, H.; Trabelsi, H.; Boujdaria, R.; Boussarsar, M.; Ouannes, L.; Bouzouita, K.; Nouira, S.; Abroug, M.F.. Dose-effect of epinephrine nebulization in asthma: A comparative study versus salbutamol. Revue de Pneumologie Clinique 1997;53(4):187-191.
**Elatrous 1997a**
Elatrous S; Elidrissi H; Trabelsi H; Boujdaria R; Boussarsar M; Ouannes L; Bouzouita K; Nouira S; Abroug MF. Dose-effect of epinephrine nebulization in asthma: a comparative study versus salbutamol.

<ORIGINAL> EFFET-DOSE DE LA NEBULISATION D'ADRENALINE DANS L'ASTHME: ETUDE COMPARATIVE AVEC LE SALBUTAMOL. Revue de pneumologie clinique 1997;53(4):187-191.

**EspinosaAyala 1980**

Espinosa Ayala, J.; Castañeda Castañeyra, E.; Martínez-Cairo, S.; Rodríguez, D.. Comparative effect of terbutaline and adrenaline in the treatment of bronchial asthma. Boletin medico del Hospital Infantil de Mexico 1980;37(3):375-381.

**Fanta 1986**

Fanta CH; Rossing TH; McFadden ER. Treatment of acute asthma. Is combination therapy with sympathomimetics and methylxanthines indicated? American journal of medicine 1986;80(1):5-10.

**Fireman 1975**

Fireman, P.; Linarelli, L.G.; Friday, G.A.. Epinephrine dose response test in severe asthma. Journal of Allergy and Clinical Immunology 1975;55(2).

**Fossati 1975**

Fossati, C.. Drug treatment of bronchial asthma. Clinica Terapeutica 1975;72(4):365-391.

**Fourton 1950**

FOURTON, A.. [Asthma and adrenalin-theophylline aerosol; emergency and ambulatory treatment]. Revue générale de médecine & de chirurgie de l'Union française 1950;27(566):2241-2246.

**Fradkin 1979**

Fradkin, I.M.; Kokin, V.S.. Emergency therapy of bronchial asthma attacks in children. Fel"dsher i akusherka 1979;44(5):43-47.

**Franchini 2010**

Franchini, S.; Marinosci, A.; Cicenia, G.. Emergency treatment of asthma. New England Journal of Medicine 2010;363(26):2567. [DOI: 10.1056/NEJMc1010476]

**Francis 1933**

Francis, A.. Asthma and Adrenaline. British Medical Journal 1933;2(3798):757. [DOI: 10.1136/bmj.2.3798.757-b]

**Frouchtman 1950**

FROUCHTMAN, R.; SEGIMON, J.. Treatment of bronchial asthma with drug aerosols.. Medicina 1950;30(612):398-401.

**Geisler 1983**

Geisler, L.S.. Epinephrine in treatment of status asthmaticus? Deutsche Medizinische Wochenschrift 1983;108(24):961.

**Glass 1973**

Glass P; Dulfano MJ. Evaluation of a new B 2 adrenergic receptor stimulant, terbutaline, in bronchial asthma. I. Subcutaneous comparison with epinephrine. Current therapeutic research, clinical and experimental 1973;15(4):141-149.

**Gotz 1981**

Gotz VP; Brandstetter RD; Mar DD. Bronchodilatory effect of subcutaneous epinephrine in acute asthma. Annals of emergency medicine 1981;10(10):518-520.

**Graeser 1935**

Graeser, J.B.; Rowe, A.H.. Inhalation of epinephrine for the relief of asthmatic symptoms. Journal of Allergy 1935;6(5):415-420. [DOI: 10.1016/S0021-8707(35)90188-5]

**Grater 1958**

Grater, W.C.; Shuey, C.B.. Medihaler in asthma: A double blind study. Southern Medical Journal 1958;51(12):1600-1602. [DOI: 10.1097/00007611-195812000-00022]

**Graterw 1958**

GRATER W C; SHUEY C B. Medihaler in asthma: a double blind study. Southern medical journal 1958;51(12):1600-1602.

**Grauvilarrubias 1951**

GRAU VILARRUBIAS, J.; MARTINEZ GAENSLY, C.. R-epinephrine aersols in the symptomatic treatment of bronchial asthma; spirometric studies.. Revista médica de Chile 1951;79(3):162-167.

**Grebe 1983**
Grebe, I. Pharmacotherapy in severe asthma attacks. Deutsche Medizinische Wochenschrift 1983;108(46):1775.
**Green 2003**
Green, S.M.. Intravenous epinephrine in asthma? A word of caution. Annals of Emergency Medicine 2003;41(5):712-713. [DOI: 10.1067/mem.2003.145]
**Hartman 1945**
HARTMAN, M.M. Parenteral use of butanefrine in asthma; a comparison with epinephrine. Annals of allergy 1945;3:366-368.
**Haughey 1952**
HAUGHEY, W. First use of epinephrine (adrenalin) in asthma.. Journal - Michigan State Medical Society 1952;51(2):216.
**Helander 1967**
Helander, S.; Lindell, S.E.; Lindholm, B.; Söderholm, B.; Westling, H. The hemodynamic and respiratory effects of adrenaline and theophylline derivatives in bronchial asthma.. Scandinavian journal of respiratory diseases 1967;48(1):45-57.
**Herraizballestro 1953**
HERRAIZ BALLESTERO, L.; RODRIGUEZ FONTELA, C.; SIMKIN, B. Treatment of bronchial asthma with associated sympathomimetics and sympatholytics drugs.. Prensa médica argentina 1953;40(3):153-155.
**Hioco 1951**
HIOCO, D.; SAMTER, M.; KARK, R.M.; BEST, W.R. The response of patients with bronchial asthma to epinephrine and to adreno-corticotropic hormone.. The Journal of clinical endocrinology and metabolism 1951;11(4):395-407. [DOI: 10.1210/jcem-11-4-395]
**Hughes 1978**
Hughes, J.A. Management of severe acute asthma. British Medical Journal 1978;1(6123):1350. [DOI: 10.1136/bmj.1.6123.1350-b]
**Hurst 1940**
Hurst, A.F. Adrenaline in Asthma. British Medical Journal 1940;2(4157):337. [DOI: 10.1136/bmj.2.4157.337-a]
**Johansen 1974**
Johansen S. Clinical comparison of intramuscular terbutaline and subcutaneous adrenaline in bronchial asthma. European journal of clinical pharmacology 1974;7(3):163-167.
John 2010
John BM; Singh D. Comparision of nebulised salbutamol and L-epinephrine in first time wheezy children. Armed forces medical journal, india 2010;66(1):9-13.
**Johnston 1963**
JOHNSTON, T.G. The treatment of bronchial asthma and emphysema. The Journal of the Arkansas Medical Society 1963;59:339-342.
**Josephson 1981**
Josephson, G.W. A controlled trial of the use of single versus combined-drug therapy in the treatment of acute episodes of asthma. American Review of Respiratory Disease 1981;124(6):765.
**Kanter 1970**
Kanter, S.D. Treatment of bronchial asthma with diadynamoelectrophoresis of adrenaline. Voprosy kurortologii, fizioterapii, i lechebnoi fizicheskoi kultury 1970;35(2):169-170.
**Karetzky 1974**
Karetzky, M.S.; Brandstetter, R.D.; Meyer, R.C.; Tedaldi, E.M.. Acute asthma. Part I: A comparison of the immediate effects of six different modes of therapy. American Journal of the Medical Sciences 1974;267(4):213-224. [DOI: 10.1097/00000441-197404000-00002]
**Karetzky 1978**
Karetzky, M.S.. Acute asthma. Part II. The effects of therapy on heart rate and blood pressure. American Journal of the Medical Sciences 1978;275(3):319-327. [DOI: 10.1097/00000441-197805000-00009]

**Karetzky 1980**
Karetzky MS. Acute asthma: the use of subcutaneous epinephrine in therapy. Annals of allergy 1980;44(1):12-14.
**Kennedy 1964**
Kennedy MC; Thursby-Pelham DC. Some adrenergic drugs and atropine methonitrate given by inhalation for asthma: a comparative study. British medical journal 1964;1(5389):1018-1021.
**Kennedy 1972**
Kennedy MC; Dash CH. The bronchodilator effect of a new adrenergic aerosol--Salmefamol. Acta allergologica 1972;27(1):22-26.
**Khaldi 1998b**
Khaldi F; Salem N. Comparison of the effect of subcutaneous adrenaline and terbutaline in infants with acute asthma.. Archives de pediatrie 1998;5(7):745-748. [DOI: 10.1016/S0929-693X%2898%2980056-0]
**Khaldi 1998c**
Khaldi F; Salem N. Comparison of the effect of subcutaneous adrenaline and terbutaline in infants with acute asthma: <ORIGINAL> COMPARAISON DE L'EFFET DE L'INJECTION SOUS-CUTANEE D'ADRENALINE ET DE TERBUTALINE DANS LA CRISE D'ASTHME DU NOURRISSON. Archives de pediatrie 1998;5(7):745-748.
**Kim 2012**
Kim, J.H.. Use of terbutaline and epinephrine in acute management of asthma. Avoiding Common Errors in the Emergency Department 2012;665.
**Kjellman 1980**
Kjellman B; Tollig H; Wettrell G. Inhalation of racemic epinephrine in children with asthma. Dose-response relation and comparison with salbutamol. Allergy 1980;35(7):605-610.
**Kobkitsumongkol 1994**
Kobkitsumongkol P. Comparison between the therapeutic effect of terbutaline dry in powder inhaler and adrenaline (subcutaneous injection) in acute mild to moderate asthmatic attack children. Songklanagarind medical journal 1994;12(4):187-188.
**Konig 1978**
Konig, P.. Treatment of severe attacks of asthma in children with nebulized B2 adrenergic agents. Annals of Allergy 1978;40(3):185-188.
**Kornberg 1992**
Kornberg, A.E.; Zuckerman, S.; Welliver, J.R.; Mezzadri, F.; Aquino, N.. Effect of injected long-acting epinephrine in addition to aerosolized albuterol in the treatment of acute asthma in children. Nursing 1992;22(5):1-3.
**Krasner 1940**
Krasner, G.. Adrenaline in Asthma. British Medical Journal 1940;2(4167):684. [DOI: 10.1136/bmj.2.4167.684]
**Krasner 1941**
Krasner, G.. Adrenaline in Asthma. British Medical Journal 1941;1(4180):255. [DOI: 10.1136/bmj.1.4180.255]
**Levy 1948**
Levy II, L.; Seabury, J.H.. Spirometric evaluation of "Ethyl-Nor-Epinephrine" in bronchial asthma. Journal of Allergy 1948;19(1):58-61. [DOI: 10.1016/0021-8707(48)90077-X]
**Lewis 1985**
Lewis, J.E.; Richards, W.; Church, J.A.; Haftel, A.; Keens, T.G.. Therapy of acute asthma: I. Evaluation of successive bronchodilator treatments. Annals of Allergy 1985;55(3):472-475.
**Limthongkul 1989**
Limthongkul, S.. Emergency room assessment and adrenaline treatment of patients with acute asthma of different severity. Journal of the Medical Association of Thailand 1989;72(6):338-345.
**Lindholm 1966**

Lindholm B; Helander E. The effect on the pulmonary ventilation of different theophylline derivatives compared to adrenaline and isoprenaline. Acta allergologica 1966;21(4):299-306.
**Lockey 1943**
Lockey, S.D.. Inhalation of oxygen and 1:100 epinephrine hydrochloride plus five per cent glycerin for the relief of asthmatic attacks. Journal of Allergy 1943;14(5):382-385. [DOI: 10.1016/S0021-8707(43)90252-7]
**Lockey 1945**
LOCKEY, S.D.. Inhalation of 10 percent carbon dioxide and 90 percent oxygen plus 1:100 glycerinized epinephrine hydrochloride for the relief of asthmatic attacks.. Annals of allergy 1945;3:362-365.
**Lopezbarrantes 1951**
LOPEZ BARRANTES, V.; LAHOZ NAVARRO, F.; RUBIALES, A.; PACIOS, A.. Effects of isopropyl adrenalin in bronchial asthma.. Revista clínica española 1951;42(6):393-397.
**Mondal 2014**
Mondal, P.; Kandala, B.; Ahrens, R.; Chesrown, S.E.; Hendeles, L.. Nonprescription Racemic Epinephrine for Asthma. Journal of Allergy and Clinical Immunology: In Practice 2014;2(5):575-578. [DOI: 10.1016/j.jaip.2014.02.014]
**Morice 1986**
Morice, A.; Sever, P.. Adrenaline, bronchoconstriction and asthma. British Medical Journal (Clinical research ed.) 1986;293(6546):539-540. [DOI: 10.1136/bmj.293.6546.539]
**Naspitz 1985**
Naspitz, C.K.; Sole, D.; Wandalsen, N.F.. Treatment of acute asthma attack. Comparison between epinephrine (s.c.) and fenoterol (by inhalation). Jornal de Pediatria 1985;58(4):173-178.
**Naspitz 1989**
Naspitz, C.K.; Sole, D.; Wandalsen, N.F.. Treatment of acute bronchial asthma. Comparison between epinephrine (subcutaneous) and fenoterol (inhalation). Revista Brasileira de Clinica e Terapeutica 1989;18(1-2):41-46.
**NCT01705964 2012**
NCT01705964. Intramuscular Epinephrine as an Adjunctive Treatment for Severe Pediatric Asthma Exacerbation. Https://clinicaltrials.gov/show/nct01705964 2012.
**Ngamphaiboon 1989**
Ngamphaiboon J; Chumdermpadetsuk S. Nebulized salbutamol vs injected epinephrine in the treatment of acute asthma in children. Chulalongkorn medical journal 1989;33(9):669-673.
**Nielsen 1936**
Nielsen, NielsA.. TREATMENT OF ASTHMATIC ATTACKS BY INHALATION OF ADRENALINE. The Lancet 1936;228(5902):848-849. [DOI: 10.1016/S0140-6736(00)48292-9]
**Noseda 1989**
Noseda, A.; Yernault, J.C.. Sympathomimetics in acute severe asthma: Inhaled or parenteral, nebulizer or spacer? European Respiratory Journal 1989;2(4):377-382.
**Nu 1971**
Nõu E. A clinical comparison of subcutaneous doses of Terbutaline and adrenaline in bronchial asthma. Scandinavian journal of respiratory diseases 1971;52(4):192-198.
**Nutbeam 2009**
Nutbeam, T.; Fergusson, A.. Endotracheal adrenaline in intubated patients with asthma. Emergency Medicine Journal 2009;26(5):359. [DOI: 10.1136/emj.2009.074088]
**Park 1941**
Park, M.. Adrenaline in Asthma. British Medical Journal 1941;1(4175):65. [DOI: 10.1136/bmj.1.4175.65-a]
**Pavlakou 2004**
Pavlakou G; Tsimboukis S; Antoniou D; Tsoukala V; Tsarouch E; Palavra M. The administration of nebulised racemic epinephrine in asthma exacerbations. European respiratory journal 2004;24(Suppl 48):541s.
**Permpikul 1990**

Permpikul C; Youngchaiyud P; Charoenratanakul S. Treatment of acute severe asthma with inhaled low dose terbutaline or subscutaneous adrenaline. Thai journal of tuberculosis and chest diseases 1990;11(4):183-191.

**Phanichyakarn 1981a**

Phanichyakarn P; Ananthachai C; Direkwattanachai C. Adrenaline and terbutaline in treatment of acute asthmatic attacks on children. Chotmaihet thangphaet [journal of the medical association of thailand] 1981;64(9):428-431.

**Phanichyakarn 1989a**

Phanichyakarn P. Comparison of subcutaneous injections of terbutaline, salbutamol and adrenaline in acute asthmatic attacks in children. Chotmaihet thangphaet [Journal of the Medical Association of Thailand] 1989;72(12):692-696.

**Pinnas 1991**

Pinnas JL; Schachtel BP; Chen TM; Roseberry HR; Thoden WR. Inhaled epinephrine and oral theophylline-ephedrine in the treatment of asthma. Journal of clinical pharmacology 1991;31(3):243-247.

**Pliss 1981**

Pliss LB; Gallagher EJ. Aerosol vs injected epinephrine in acute asthma. Annals of emergency medicine 1981;10(7):353-355.

**Rees 1967**

Rees HA; Millar JS; Donald KW. Adrenaline in bronchial asthma. Lancet (london, england) 1967;2(7527):1164-1167.

**Refsum 1956**

REFSUM, H.E. Treatment of severe attacks of asthma with continuous intravenous administration of adrenaline.. Nordisk medicin 1956;55(8):257-259.

**Riding 1970**

Riding WD; Dinda P; Chatterjee SS. The bronchodilator and cardiac effects of five pressure-packed aerosols in asthma. British journal of diseases of the chest 1970;64(1):37-45.

**Rohr 1986**

Rohr AS; Spector SL; Rachelefsky GS; Katz RM; Siegel SC. Efficacy of parenteral albuterol in the treatment of asthma. Comparison of its metabolic side effects with subcutaneous epinephrine. Chest 1986;89(3):348-351.

**Ross 1946**

Ross, J.. Intravenous adrenaline in asthma. British Medical Journal 1946;2(4467):242. [DOI: 10.1136/bmj.2.4467.242]

**Rossing 1980**

Rossing TH; Fanta CH; Goldstein DH; Snapper JR; McFadden ER. Emergency therapy of asthma: comparison of the acute effects of parenteral and inhaled sympathomimetics and infused aminophylline. American review of respiratory disease 1980;122(3):365-371. [DOI: 10.1164/arrd.1980.122.3.365]

**Rossing 1981**

Rossing TH; Fanta CH; McFadden ER. A controlled trial of the use of single versus combined-drug therapy in the treatment of acute episodes of asthma. American review of respiratory disease 1981;123(2):190-194. [DOI: 10.1164/arrd.1981.123.2.190]

**Royle 1938**

Royle, H.; Hutchinson, C.A.. Adrenaline Treatment of Asthma. British Medical Journal 1938;2(4064):1112. [DOI: 10.1136/bmj.2.4064.1112-b]

**Schwartz 1980a**

Schwartz AL; Lipton JM; Warburton D; Johnson LB; Twarog FJ. Management of acute asthma in childhood. A randomized evaluation of beta-adrenergic agents. American journal of diseases of children (1960) 1980;134(5):474-478.

**Sharma 1985**

Sharma TN; Kala D; Gupta PR; Purohit SD; Gupta RB; Sisodia RS. Comparison of subcutaneous salbutamol and terbutaline with adrenaline. Indian journal of chest diseases & allied sciences 1985;27(1):27-30.

**Sly 1969**

Sly, R.M.. Comparison of Bronkephrine and epinephrine in the treatment of acute asthmatic attacks in children.. Annals of Allergy 1969;27(8):384-389.

**Snider 1955**

SNIDER, G.L.; BARNETT, K.; RADNER, D.B.; MOSKO, M.M.. The evaluation of bronchodilator drugs in the treatment of asthma.. The Journal of laboratory and clinical medicine 1955;46(3):348-358.

**Streeton 1977**

Streeton, J.A.; Tashkin, D.P.. Epinephrine vs newer bronchodilator drugs in the treatment of acute asthma. Chest 1977;72(6):801. [DOI: 10.1378/chest.72.6.801a]

**Taub 1968**

Taub, S.J.. Effects of epinephrine on asthmatic children.. Eye, ear, nose & throat monthly 1968;47(5):256-257.

**Taub 1971**

Taub, S.J.. Drug therapy of the asthmatic child.. Eye, ear, nose & throat monthly 1971;50(5):186-187.

**Teoh 1979**

Teoh PC. Clinical evaluation of intravenous hexoprenaline in bronchial asthma. Annals of the academy of medicine (singapore) 1979;8(2):144-147.

**Ting 1991**

Ting CK; Liao MH. A comparative study of epinephrine injection and beta 2-agonist inhalation in the treatment of childhood asthma. Zhonghua minguo xiao ER ke yi xue hui za zhi [journal]. Zhonghua minguo xiao ER ke yi xue hui 1991;32(6):372-381.

**Tirot 1992**

Tirot, P.; Bouachour, G.; Varache, N.; Harry, P.; Bourrier, P.; Chennebault, J.M.; Alquier, P.. The use of intravenous adrenaline in acute severe asthma. Revue des Maladies Respiratoires 1992;9(3):319-323.

**Toivonen 1964**

TOIVONEN, S.; TARPILA, S.; BACKMAN, A. SPIROMETRIC STUDIES OF THE EFFECT OF 2 ADRENALINE DERIVATIVES ON BRONCHIAL ASTHMA.. Nordisk medicin 1964;72:1047-1049.

**Turner 1938**

Turner, H.M.S.; Francis, A.. Adrenaline Treatment of Asthma. British Medical Journal 1938;2(4065):1178. [DOI: 10.1136/bmj.2.4065.1178-a]

**Turpeinen 1983**

Turpeinen M; Kuokkanen J; Backman A. Adrenaline and salbutamole in acute asthma of children. Suomen lãµãµkãµrilehti 1983;38(8):652-654.

**VonHundelshausen 1983**

Von Hundelshausen, B.; Tempel, G.; Schneck, H.J.. Epinephrine in the treatment of status asthmaticus. Deutsche Medizinische Wochenschrift 1983;108(19):760-761.

**Weber 1968**

Weber, J. Treatment of acute asthmatic crisis. Praxis 1968;57(2):41-43.

**Weinberger 1974**

Weinberger MM; Bronsky EA. Evaluation of oral bronchodilator therapy in asthmatic children. Bronchodilators in asthmatic children. Journal of pediatrics 1974;84(3):421-427.

**Zeggwagh 2002a**

Zeggwagh, A.A.; Abouqal, R.; Madani, N.; Abidi, K.; Moussaoui, R.; Zekraoui, A.; Kerkeb, O.. Compared efficacy of nebulized adrenaline and salbutamol in acute severe asthma. A randomized, prospective and controlled study. Annales Francaises d'Anesthesie et de Reanimation 2002;21(9):703-709. [DOI: 10.1016/S0750-7658(02)00779-7]
